# A Tutorial on Discrete Event Simulation Models in R using a Cost-Effectiveness Analysis Example

**DOI:** 10.1101/2025.05.15.25327635

**Authors:** Mauricio Lopez-Mendez, Jeremy D. Goldhaber-Fiebert, Fernando Alarid-Escudero

## Abstract

Discrete Event Simulation (DES) is a flexible and computationally efficient approach for modeling diverse processes; however, DES remains underutilized in healthcare and medical decision-making due to a lack of reliable and reproducible implementations. We developed an open-source DES framework in R to simulate individual-level state-transition models (iSTMs) in continuous time accounting for treatment effects, time dependence on state residence, and age-dependent mortality. Our DES implementation employs a modular and easily adaptable structure, with each module corresponding to a unique transition between health states. To simulate the evolution of the process (i.e., individual state-transitions), we adapted the next-reaction algorithm from the stochastic chemical reactions literature. Simulation-time dependence (age-dependent mortality) and state residence time dependence (transition from Sick to Sicker) are seamlessly incorporated into the DES framework via validated non-parametric and parametric sampling routines (e.g., inversion method) of event times. Treatment effects are integrated as scaling factors of the hazard functions (proportional hazards). We illustrate the framework’s benefits by implementing the Sick-Sicker Model and conduct a cost-effectiveness analysis and probabilistic analysis. We also obtain epidemiological outcomes of interest from the DES output, such as disease prevalence, survival probabilities, and distributions of state-specific dwell times. Our DES framework offers a reliable and accessible alternative that enables the simulation of more realistic dynamics of state-transition processes at potentially lower implementation and computational costs than discrete-time iSTMs.

**Higlights:**

- Discrete event simulation (DES) is a flexible and efficient approach to simulate diverse processes in model-based decision analysis.
- The tutorial presents an open-source DES framework in R to simulate individual-level State-Transition Models (iSTMs) in continuous time.
- The modular structure of our DES framework accommodates treatment effects, time-dependent transitions, and age-dependent mortality using validated sampling methods.
- The coded example uses the Sick-Sicker model to compute a cost-effectiveness analysis (CEA), epidemiological outcomes, probabilistic analysis (PA), and value of information analysis (VOI).

## 1. Introduction

Discrete event simulation (DES) is a general framework for simulating dynamic systems stochastically [1, 2, 3] . The salient feature is that the variables of interest are represented by “events” and the system’s state is only updated when events occur. The DES framework can represent dynamic processes that evolve over continuous or discrete time, and can include multiple sources of within- and between-person variation[4], as well as different types of dependencies on time and previous states of the system (i.e., event history).

DES models apply to many situations as they are highly general. They are commonly used to analyze queuing systems, and more recently, they have been applied to model epidemiological and clinical phenomena, such as disease progression and transmission (e.g., [5, 6]. They are particularly valuable for modeling complex dynamics that can inform medical, pharmacological, and healthcare decision-making (e.g., [7, 8, 9]).

One advantage of DES is its computational and memory efficiency, even when representing systems involving large numbers of heterogeneous individuals facing complex dynamics. Discrete-time individual state-transition simulation models (iSTMs) (i.e., microsimulations) are frequently used to model systems with complex dynamics and heterogeneity. However, such models can require large amounts of computational power to generate sufficient simulated trajectories to make stable estimates of the key outcomes. This is because each trajectory in an iSTM requires evaluation of potential transitions at fixed time intervals, generating a pseudorandom number for each transition in each interval for each trajectory. However, the evolution of trajectories in a DES is “event-driven”, meaning that evaluation is only necessary as events occur, greatly reducing the computation and memory requirements.

This tutorial aims to present a modular DES framework for model-based CEA and motivate its use with a healthcare example. Our framework emphasizes modeling principles that support a general implementation of DES that readers will find helpful for building their own implementation from first principles and without necessarily depending on specialized software. First, we provide a theoretical description of the DES framework. Second, we illustrate how to simulate time-dependent state-transition models (STM) using DES. Third, we conceptualize how to include treatment effects as well as covariates that can potentially be time-varying. DES yields equivalent outputs to those from cohort STMs and iSTMs that can be used in cost-effectiveness analysis (CEA) and health-technology assessments (HTAs). Finally, we provide a modular example in the R programming language [10] with the code to generate economic and epidemiological outcomes of interest in CEA and HTA, which can be adapted to more complex iSTMs following our DES framework. Readers will find that DES can be used to implement models in a straightforward manner similar to their prior experience with other techniques while enabling them to access its advantages without substantial additional programming efforts.

## 2. Theoretical Framework and Notation

A stochastic process is a collection of random variables, typically indexed by time. The domain of these random variables can be continuous or discrete [2].Mathematically, a DES corresponds to a continuous-time stochastic process over a discrete, finite event space. Compared to discrete-time STMs [11, 12, 13], continuous-time STMs require a different set of mathematical tools to analyze the simulated individuals’ dynamics and behaviors.

### 2.1 Event-driven Dynamics in an STM

The evolution of a stochastic process with a discrete, finite state space can be described by the occurrence of “events” over time, defined by three components in an STM:

- An “event” is a change of state, i.e., a change in the discrete values of a variable *X*_*t*_, and the set of all possible events is denoted as the “event-space”. In the context of STMs, we can think of the set of unique transitions between states as the set of all possible events.
- The random variable *T*_*n*_ denotes the *n*^*th*^ event time (*n* = 1,2, … < ∞), i.e., the time at which the *n*^*th*^ change of state occurs. An example of an event time is the time at which a patient transitions from healthy to sick.
- Inter-event time (i.e., dwell time), denotes the time spent in a given state before transitioning to another state, and is described mathematically as the time increment between consecutive events, *τ*_*n* − 1_ = (*T*_*n*_ − *T*_*n* − 1_).

Since (*T*_*n*_, *T*_*n* − 1_) are random variables, the inter-event time, *τ*_*n* − 1_, is a random variable with an associated cumulative distribution function (CDF), *F*(*τ*)_*r,s*_, where (*r*) indexes the origin state (value of *X*_*t*_ immediately before the *n*^*th*^ event) and (*s*) indexes the destination state (value of *X*_*t*_ after the *n*^*th*^ event).

Figure 1 illustrates these concepts for a simple three-state iSTM for one simulated person that starts in state *S*_1_ at time zero, *T*_0_ and can transition between *S*_1_ and *S*_2_, and is also at risk of transitioning to the absorbing state *S*_3_, i.e., once reached, individuals remain in *S*_3_. Each transition is an event, and the four unique transitions in the state-transition diagram represent the event-space. A realization of a single trajectory of the 3-state model can be represented either as a sequence of states visited over time, or as a sequence of events occurring over time. In a DES, we focus on the latter, and keep track of the time points when the events occur. Consider the following realization of a single trajectory: {*S*_1_, *S*_2_, *S*_1_, *S*_3_}. Discrete-time STMs would evaluate the state a person occupies at all points on a finite grid of time points, requiring evaluations *X*_*t*=0_, *X*_*t*=1_, …, *X*_*t*=*k*_; *k* < ∞. Equivalently, we can encode the same information as a sequence of events, i.e., transitions between states: (*S*_1_ → *S*_2_, *S*_2_ → *S*_1_, *S*_1_ → *S*_3_). With this event-driven representation, we only need to keep track of the three times at which the events occur (*T*_1_, *T*_2_, *T*_3_); for which we only need three evaluations 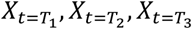.

**Figure 1.**
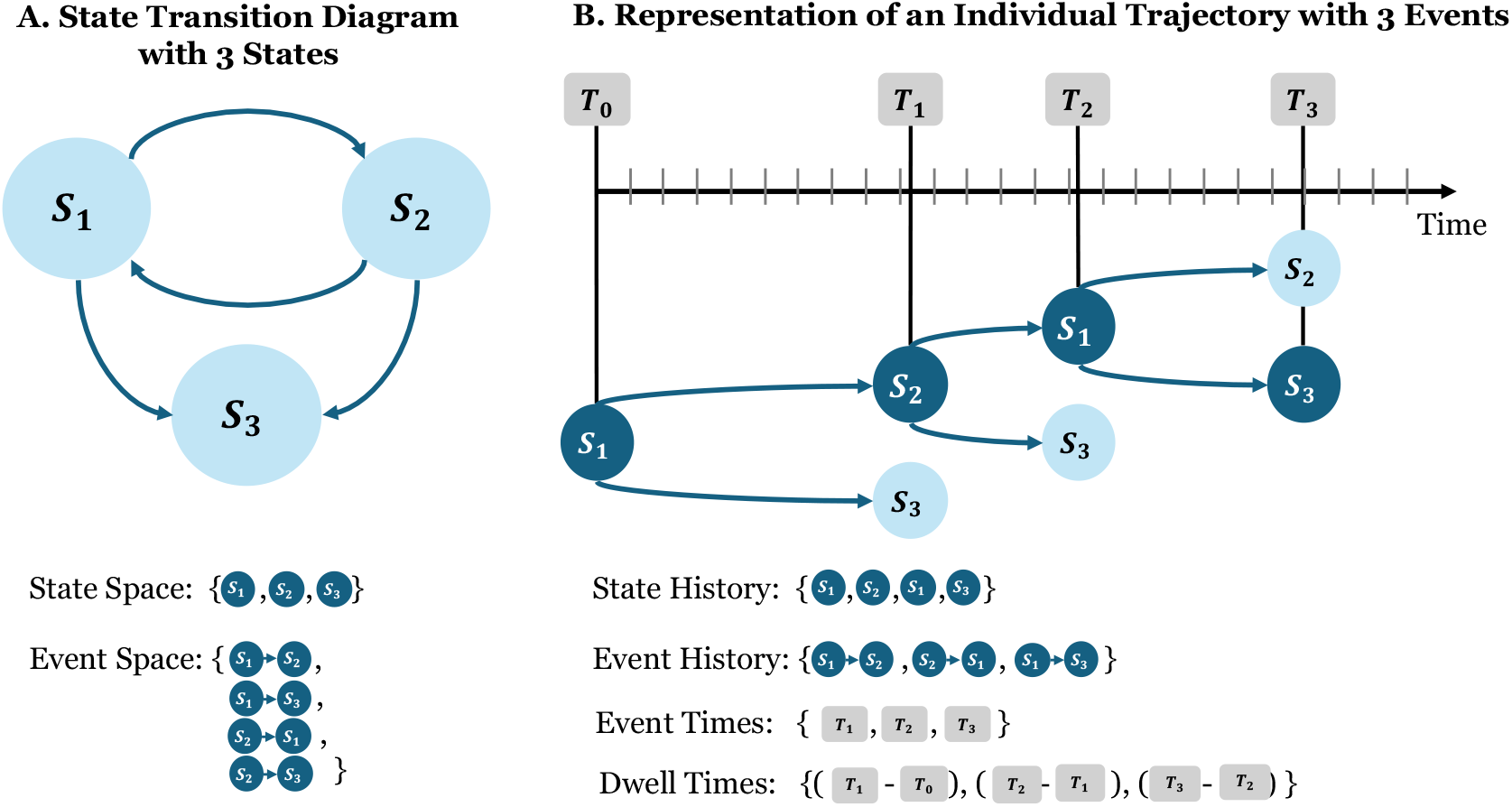
State-Transition Diagram of a Model with three States and Example of an Individual Trajectory of three Events. Panel A. State-transition diagram with three states. State space refers to the set of mutually exclusive and collectively exhaustive states that a simulated person can visit during the simulation. Event space refers to the set of unique transitions between pairs of states that a simulated person can experience during the simulation. Panel B. Representation of an individual trajectory involving the occurrence of three events. State history refers to the previosuly occupied health states. Event history refers to the previosly experienced transitions between health states. Event times are the time points at which transitions between health states occured. Dwell times refer to the elapsed time in a given state before transitioning to another state.

### 2.2 Intensity Functions and Cumulative Intensity Functions

A key concept to describe state-transition processes in continuous time is the notion of transition-specific intensity functions (also referred to as hazard functions in survival analysis)[14]. In the context of STMs, the intensity function describes the rate or instantaneous probability at which a person would transition from a state *r*, to a state *s, r* ≠ *s* [15, 16].

Let 𝕊 denote the set of possible states that a person can visit throughout the simulation time-horizon, e.g., lifetime.

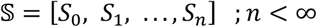

Let *X*_*t*_ ∈ 𝕊, *t* ≤ 0, be a collection of random variables denoting the state at time *t* that can only take values from the set 𝕊. For example, consider an individual at time *t* who is in state *X*_*t*_ = *r, t ≥* 0, then, the intensity of transitioning to the next state, *X*_*t* + *δ*_ = *s*, is represented by the intensity function *λ*_*r,s*_(*t*, ***z***(*t*), *ℋ*_*t*_):

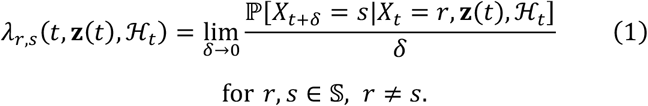

Where *λ*_*r,s*_ may depend on time *t*, covariates ***z***(*t*), and the history of states visited before the time of the current event *T*_*n*_,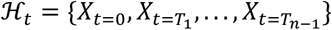. Additionally, let

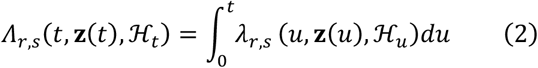

denote the cumulative intensity function that describes the accumulated rate of transition between state *r* and state *s* up to time *t*.

## 3. Evolution of a Continuous Time Process with Discrete and Finite Event-space (Simulating Sequences of Events)

### 3.1 Inter-event Time Sampling

If the process of transitioning between state *r* to state *s* follows a constant intensity function, the CDF of inter-event times *F*(*τ*)_*r,s*_ follows an exponential distribution. This corresponds to a homogeneous Poisson point process (HPPP) and has an analytic solution. However, if the intensity function depends on time, e.g., the time elapsed in state *r*, the distribution *F*(*τ*)_*r,s*_ could correspond to a non-homogeneous Poisson point process (NHPPP), which does not necessarily have a known analytic solution.

To simulate state-transition processes with an event-driven approach, validated routines are recommended for sampling the inter-event times, especially when the intensity function depends on time. In our DES implementation, we provide submodules for sampling inter-event times following either the inversion method(e.g., [17, 18] ) or the non-parametric approach [19].

### 3.2 Simulation Algorithm (Next-reaction Method)

There are at least two approaches to simulate realizations of the stochastic process underlying a DES of an iSTM in continuous time [20]. The first approach relies on choosing the transition that occurs at a faster rate immediately after an event occurs, also referred to as “next reaction” methods [21]or as an ‘event-specific distribution’ approach [22]. These algorithms require knowledge of the set of transition-specific intensity functions for each state, which allows sampling the latent times for each competing transition to determine the next event. The second approach consists of first determining the time of the next event of any type and then, conditional on that time, determining the type of event that will occur. Our DES implementation is based specifically on Anderson’s modified next reaction method [23] and adaptated for general non-exponential inter-event times [24]. This approach involves setting the transition-specific cumulative intensity functions, their corresponding inverse functions, and the adjacency matrix *A* that encodes the set of directly connected states, as represented by the state-transition diagram (i.e., for each state, determine which direct transitions the model permits).

At a given event time, *T*_*n*_, in order to determine the next transition for each person, we first determine the set of possible states that a person can transition to given their current state 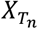. This can be done using an adjacency matrix *A*. In the example with three states (Figure 1), one person starts at S1 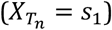 and faces two possible transitions, to S2 or to S3. Second, we sample a set of latent inter-event times 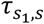, for *s* ∈ {*s*_2_, *s*_3_} via the inversion or non-parametric methods (see Section 3.1). With samples of the time from S1 to S2,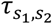, and from S1 to S3, 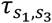, we can determine the first event out of S1. We compute the minimum inter-arrival time 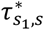 among the latent inter-event times, i.e.:

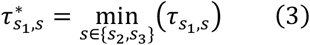

With the realized inter-event time 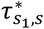, we update the current state determined by its second index *s*^*^.

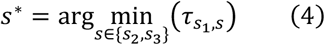

Finally, the simulation time is updated to 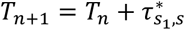, and the state at the next event time is updated to 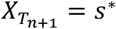.

Figure 2 illustrates these steps for the more general case of one person in a model with *m* states, starting from state S1 and facing (*m*−1) possible transitions out of S1.

**Figure 2.**
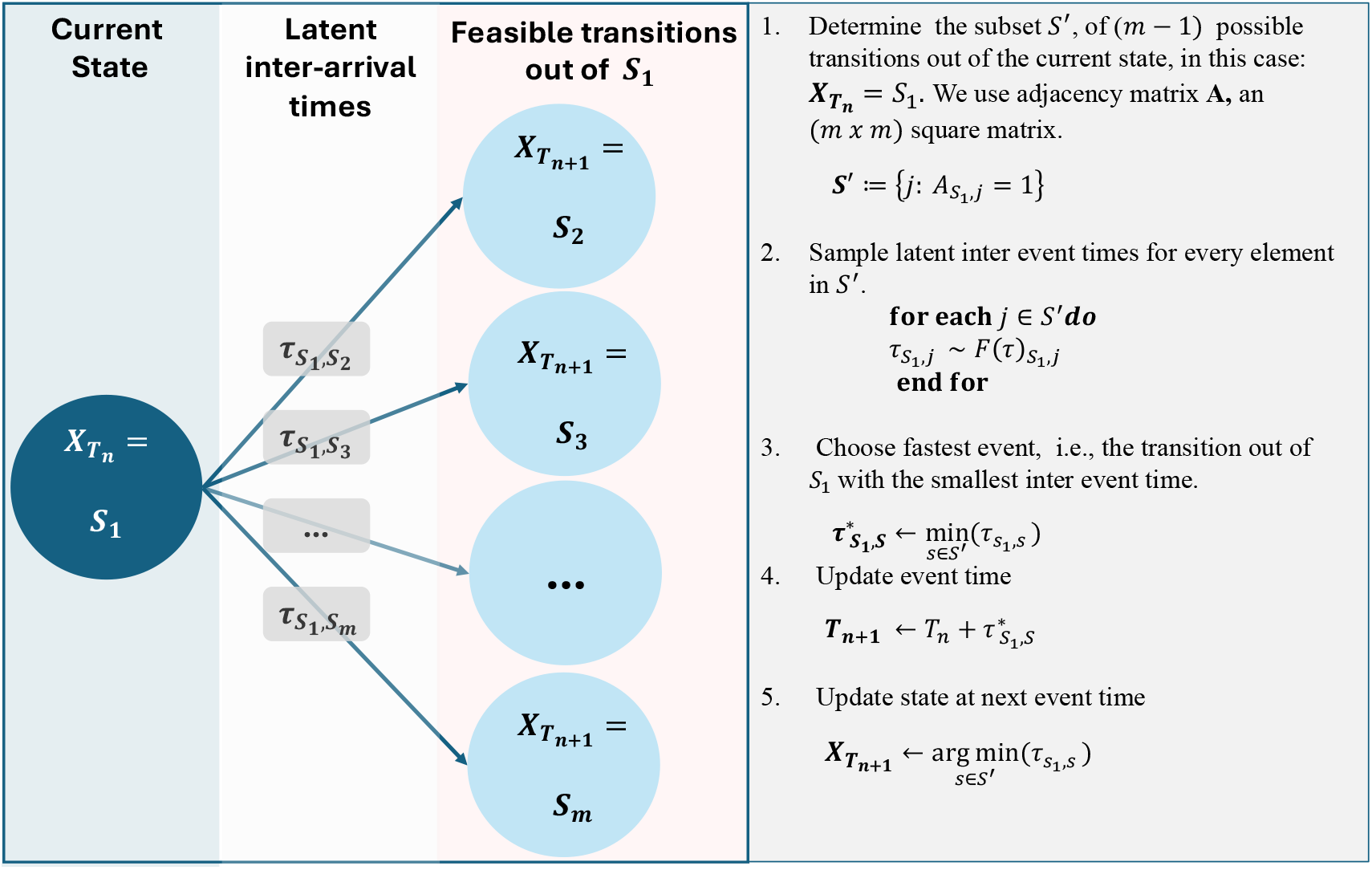
Schematic and Peseudocode of Simulation Algorithm Left. Diagram of (*m*−1) feasible transitions out of *S*_1_. Right. Pseudocode for determining the next transition and the inter-event time following the next reaction method.

**Figure 3.**
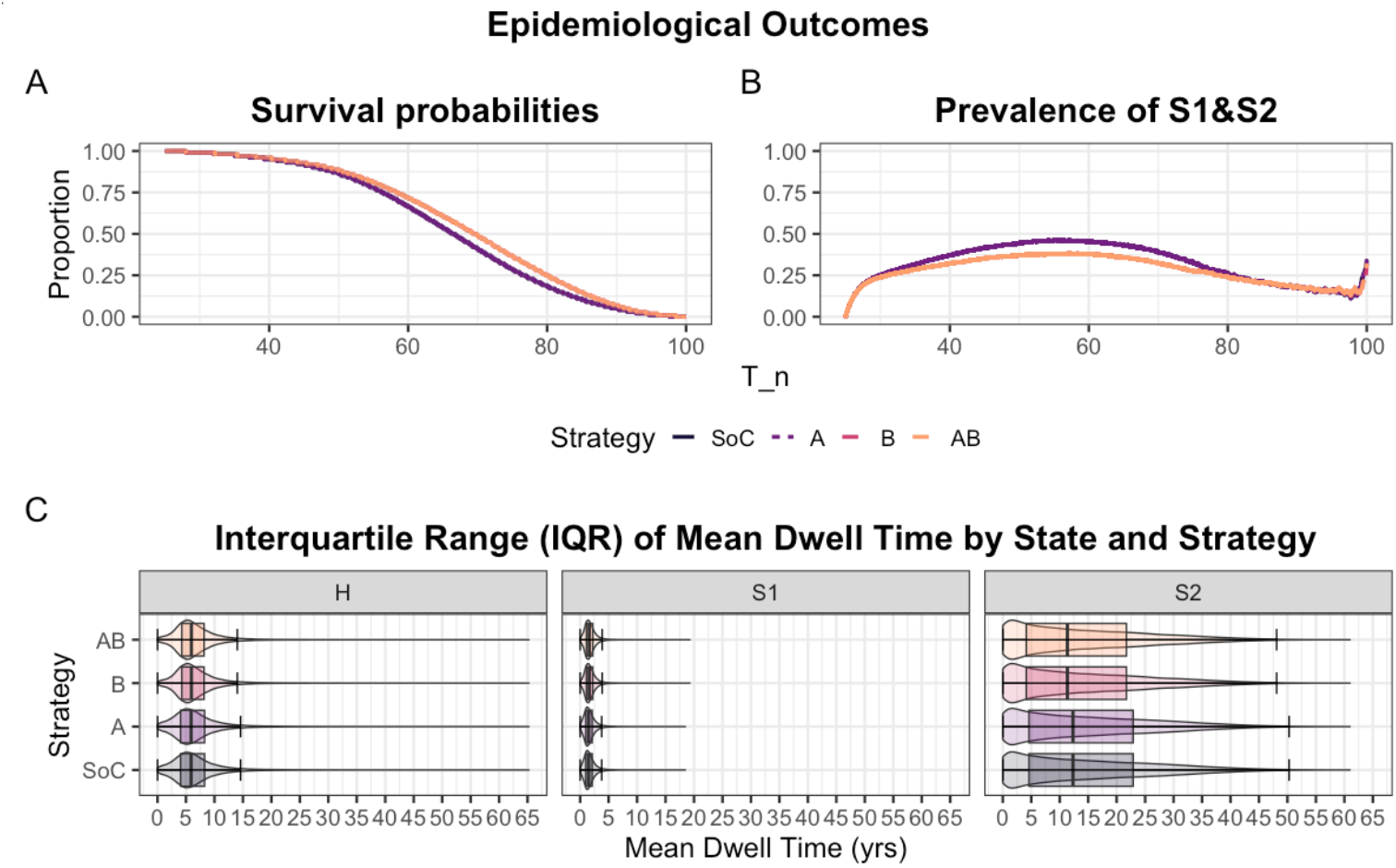
Epidemiological Outcomes Panel A.Comparison of survival probabilities under each Strategy. Strategies “SoC” and “A” do not affect transition rates hence probabilities of surviving up to time *t* are unaffected. Strategies “B”and “AB” reduce the rate of transition from Sick to Sicker states increasing the probabilities of surviving up to time *t*. Panel B. Comparison of prevalence curves under each strategy. Strategies “SoC” and “A” do not affect transition rates hence probabilities of being in a Sick state (Sick or Sicker) at time *t* are unaffected. Strategies “B” and “AB” affect transition from Sick to Sicker states reducing the probabilities in a Sick state (Sick or Sicker) at time *t*. Panel C. Comparison of the interquartile range of the distribution of individual mean dwell times in each alive state under each strategy . Strategies “B” “and”AB” reduce transition from Sick to Sicker states reducing the individual median dwell time in the Sicker state compared to strategies “SoC” and “A” that do not affect transitions between health states.

## 4. Adding Complexity to a DES

The next-reaction method relies on transition-specific intensity functions, capturing time-dependence, treatment effects and individual-level heterogeneity influencing transition rates. These complexities would require specialized techniques in a cSTM, like tunnel states, multidimensional transition probability arrays, and stratified health states [11, 12]. In contrast, DES does not require such techniques, and can incorporate treatment, covariates, time dependence, and state-history directly via transition-specific intensity functions, *λ*(*t*)_*r,s*_.

### 4.1 Time Dependence

When intensities depend on simulation time or time since state-residency started, the distribution of time in state *r* before transitioning to state *s, F*(*τ*)_*r,s*_, may be unknown analytically. However, the inversion [18] and non-parametric methods [19] offer reliable solutions for sampling from an unknown *F*(*τ*)_*r,s*_ via the cumulative intensity function *Λ*(*t*)_*r,s*_.

### 4.2 Treatment and Covariate Effects

The effects of treatments and covariates on transition-specific intensities can be modeled using the proportional hazards assumption, which provides an interpretable way to encode these effects, like hazard ratios in clinical trials. Other formulations, such as accelerated failure time, are also possible.

To illustrate how treatment and covariate effects influence transition rates under proportional hazards, consider a treatment that only affects the transition rate from state *r* to *s*. The intensity function for the *i*^*th*^ individual describing the transition from state *r* to *s* is then expressed as:

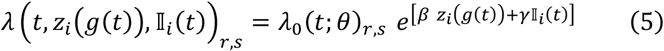

Where *λ*_0_(*t*; *θ*)_*r,s*_ is the baseline intensity function which in our framework can belong to any flexible parametric model parameterized by the vector (*θ*) (note that non-parametric specifications of *λ*_0_(*t*)_*r,s*_ are also possible); (*β*) is a vector of time-fixed covariate effects; *z*(*g*(*t*)) is a vector of covariates (possibly time-varying) and *g*(*t*) is a function of time; (*γ*) is a time-fixed scalar treatment effect; and 𝕀_*i*_ (*t*) is an indicator function of receiving the treatment, possibly time-varying. Note that this formulation makes treatment effects enter the intensity function like a covariate.

### 4.3 State-history Dependence

If the simulation stores previous simulated events, the information of previously visited states, *ℋ*_*𝓉*_, can enter the intensity function as a covariate as well; as shown in Equation 5.

### 4.4 Capacity and Resource Constraints

Depending on the application, it may be necessary to explicitly model delays due to resource or capacity constraints. Queues are well-studied models that represent continuous-time processes, compatible with DES [25, 26]. In healthcare, queues can represent capacity constraints – for instance, limited appointments for diagnostic imaging can delay diagnosis and treatment, allowing disease severity to progress.

## 5. Case study: a Cost-effectiveness Analysis using a Time-dependent Sick-Sicker model

We demonstrate the implementation of our DES framework by simulating in continuous time the time-dependent 4-state Sick-Sicker model [11, 12, 13, 27].

A cohort of 25-year-olds in the “Healthy” state (denoted “H”) is simulated until they reach age 100. Cohort members face an age-specific background mortality rate, *λ*(*t*)_*H,G*_ (v_r_HDage). Healthy individuals are also at risk of a hypothetical disease with 2 stages, “Sick” and “Sicker”, denoted by “S1” and “S2”, respectively. The disease process is described as follows:

- Healthy individuals face a constant annual rate of getting sick, *λ*_*H,S*1_ (r_HS1 in R notation). The time from H to S1, *τ*_*H,S*1_, is exponentially distributed, *τ*_*H,S*1_ *∼ Exp*(*λ*_*H,S*1_).
- Sick individuals are at risk of progressing to the more severe Sicker health state, as a function of time spent in the Sick state following a Weibull intensity parameterized as proportional hazards, i.e.,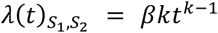, where *t* is the time spent in S1, and *β* (r_S1S2_scale) and *k* (r_S1S2_shape) are the scale and shape parameters for the Weibull hazard function, respectively. The time from S1 to S2, *τ*_*S*1,*S*2_ *∼ W eibull*(*β, k*). Both *β* and *k* are greater than zero. Sick individuals can recover and become Healthy again at a constant rate *λ*_*S*1,*H*_ (r_S1H), i.e., *τ*_*S*1,*H*_ *∼ ExpSλ*_*S*1,*H*_*T*. However, once individuals reach S2, they cannot recover; i.e., the transition rate to S1 or H from S2 is zero.
- Individuals in S1 and S2 face an increased hazard of death compared with healthy individuals, which is modeled as a relative increase to the age-specific background mortality rate with a hazard ratio (HR) of 3 (hr_S1) and 10 (hr_S2), respectively.

Individuals in S1 and S2 also experience increased health care costs and reduced quality of life (QoL) compared with individuals in H. When individuals die, they transition to the absorbing “Dead” state (denoted by “D”), with no further transitions. We use the model to compute the cohort’s expected costs and quality-adjusted life-years (QALYs) over their lifetime. We discount costs and QALYs at a 3% annual rate.

We assess the cost-effectiveness of 4 strategies: standard of care (strategy SoC); strategy A; strategy B; and a combination of strategies A and B (strategy AB).

- Strategy A administers treatment A, increasing the QoL of individuals in S1 from 0.75 (utility without treatment, u_S1) to 0.95 (utility with treatment A, u_trtA). Treatment A costs $12,000 per year (c_trtA). This strategy does not affect the QoL of individuals in S2, nor changes the risk of becoming sick or progressing through the sick states.
- Strategy B implements treatment B, reducingthe rate of progressing to Sicker by 40%, costs $13,000 per year (c_trtB), and does not affect QoL. The effect of strategy B is encoded as a hazard ratio *HR* = 0.6 (hr_S1S2_trtB).
- Strategy AB involves administering both treatments A and B with both effects and additive costs.

We assume that it is not possible to distinguish between Sick and Sicker patients; therefore, individuals in both disease states receive treatment(s) under all treatment strategies. After comparing the 4 strategies in terms of expected QALYs and costs, we calculate the incremental cost per QALY gained for non-dominated strategies.

Model parameters and the corresponding R variable names are presented in Table 1 . We follow the notation conventions described in the DARTH coding framework [28]. These conventions are proposed for clarity and consistency; analysts are welcome to adopt their own coding and naming and adjust the accompanying R code accordingly. Our implementation mainly relies on the data.table R package for memory efficiency and computational speed for handling large data sets [29]. In the appendix, we show execution time distributions of our code and describe package dependencies.

**Table 1:**
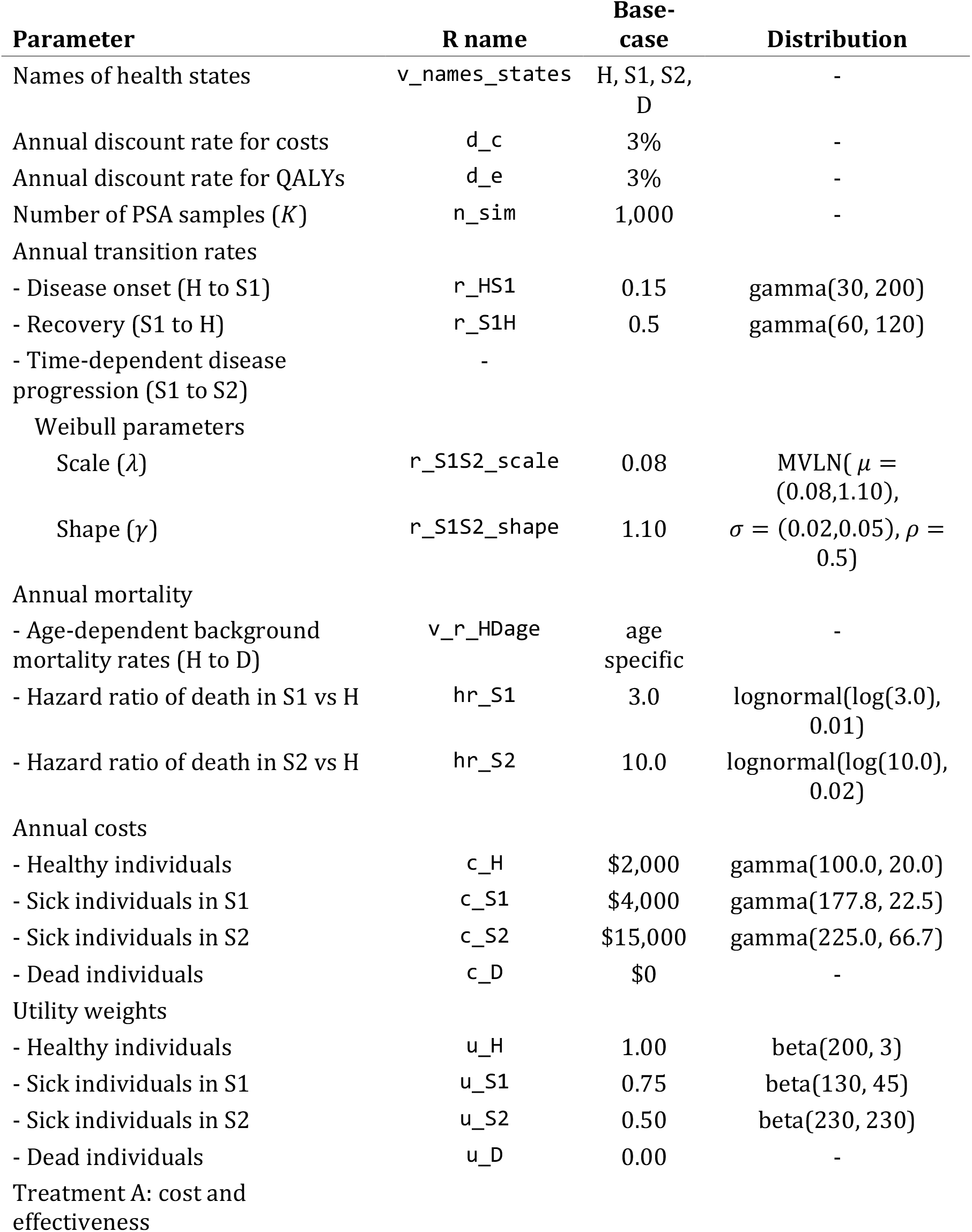
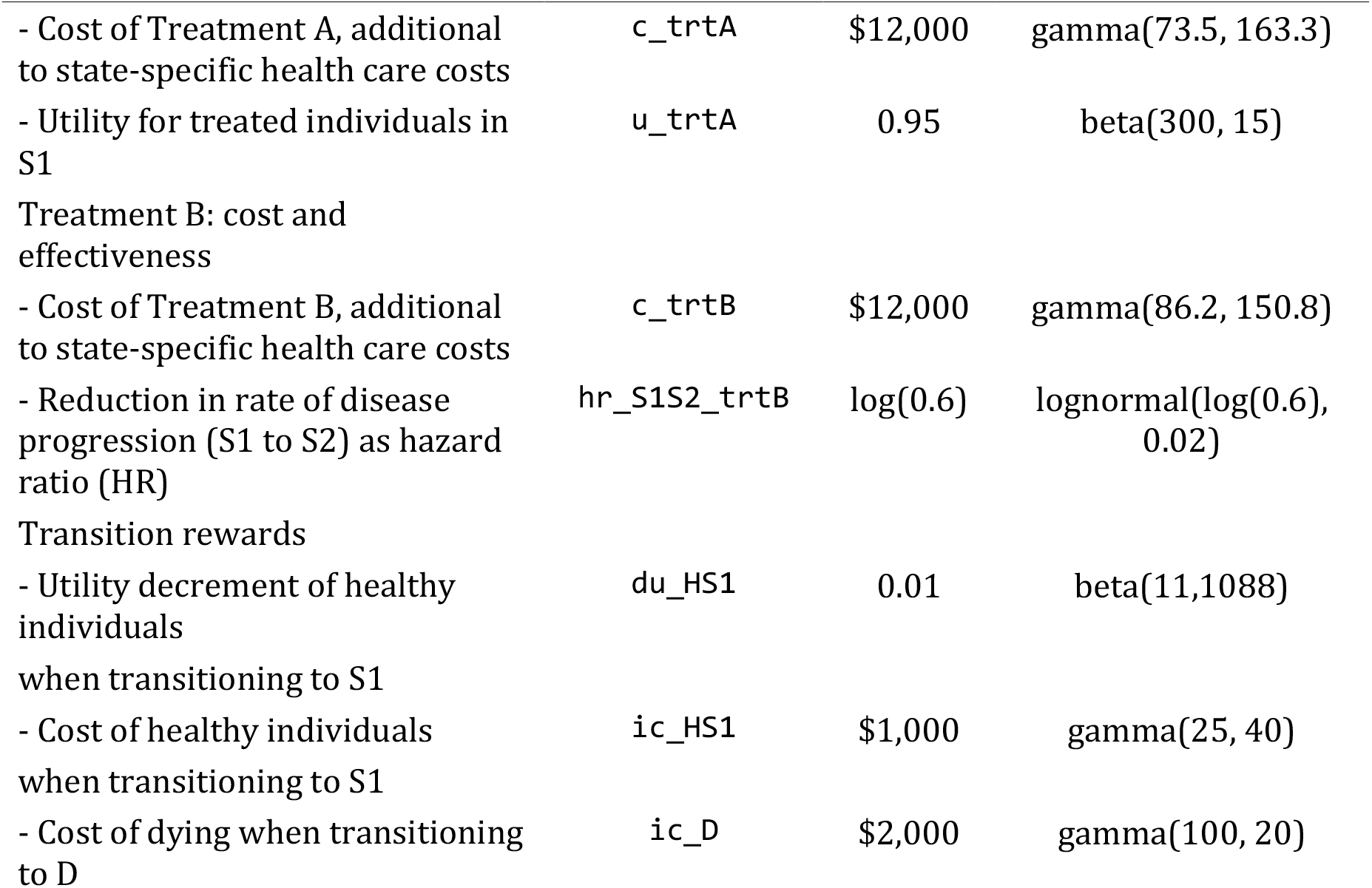
Description of parameters, their R variable name, base-case value and distribution.

### 5.2 Useful Data Objects

Our simulation framework uses three distinct data.table (dt) objects:

a. `dt_baseline`: stores baseline characteristics, initial states, and initial times for all simulated people. This dt is only needed to initialize the simulation.
b. `dt_next_event`: stores one instance of the latent transitions that can occur for each simulated person at a given time. This dt will be overwritten several times as new events occur, only storing relevant information to determine the next event for each person in the simulation.
c. `dt_event_history`: stores the history for each simulated person’s transitions and the times they occurred. This dt stores all instances of `dt_next_event` in long format.

Additionally, a numbered adjacency matrix, *A* (labeled as `m_num_adj`), is used to determine which are the possible transitions from each state. For the Sick-Sicker model the adjacency matrix is informally defined as:

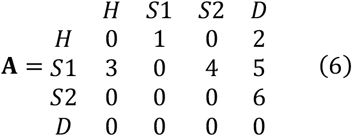

A conventional adjacency matrix is a square matrix with only 0 or 1 values, where 1 indicates a possible transition (i.e., the states are connected in the state-transition diagram), and 0 otherwise. We enumerate nonzero entries to easily identify health state transitions. Alternative implementation styles in DES often include, “event-oriented” or “process-oriented” approaches. This tutorial focuses on the continuous-time stochastic process underlying an iSTM and organizes the code around the state-transition structure defined by the adjacency matrix.

### 5.3 Sequences of Events and Event Times

The provided code generates the sequence and timing of events for each person in two modules (See eFigure 1 in the appendix).

#### 5.3.1 Module 1: Currently Occupied States

The first module reads `dt_main`, supplied as `dt_baseline` or `dt_next_event`, identifies the occupied states by simulated individuals, and creates subsets for each occupied state stored in a `list l_subsets_dt_main`. For example, if 50% start the simulation Healthy and 50% in the Sick state, the module splits `dt_baseline` into two `dt_subsets`, one with people in the Healthy state and one with people in the Sick state. It also reads the life table, dt_bckgrd_mortality and produces sick-state-specific life tables using the provided hazard ratios, `dt_mortality_Sick` and `dt_mortality_Sicker`.

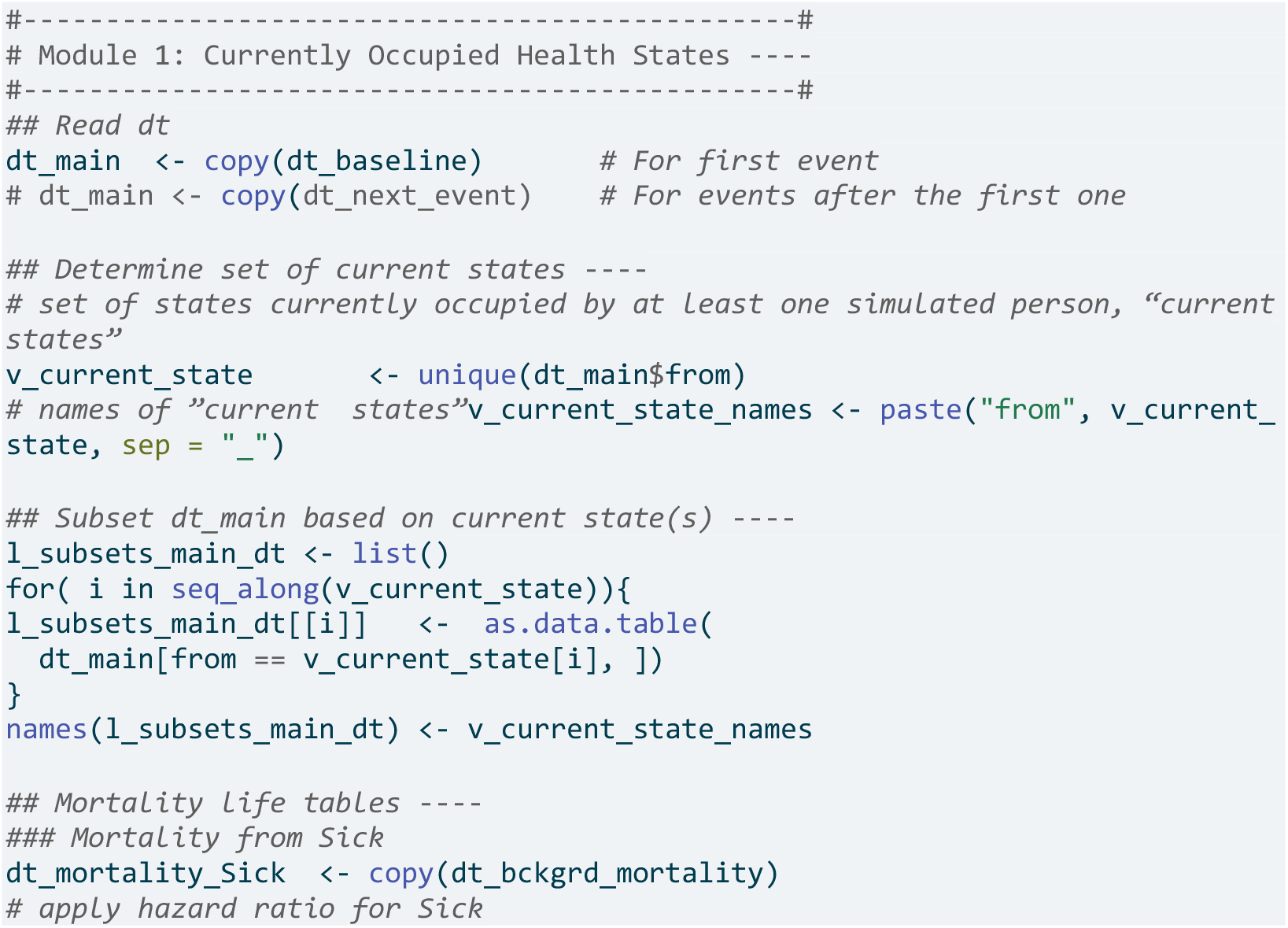
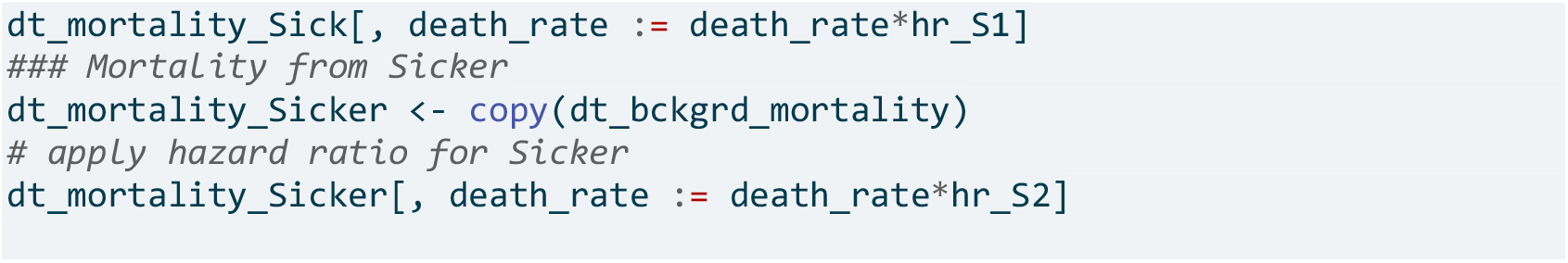

#### 5.3.2 Module 2: Transitions between Health States

This module is the core of the simulation, divided into submodules for each non-absorbing state. The Sick-Sicker model has three submodules, corresponding to the Healthy, Sick and Sicker states. Adding more submodules requires minimal extra work since submodules are standardized with four tasks, even in models with a larger state-space. The tasks for the *i*^*th*^ submodule are summarized in Table 2.

**Table 2:**
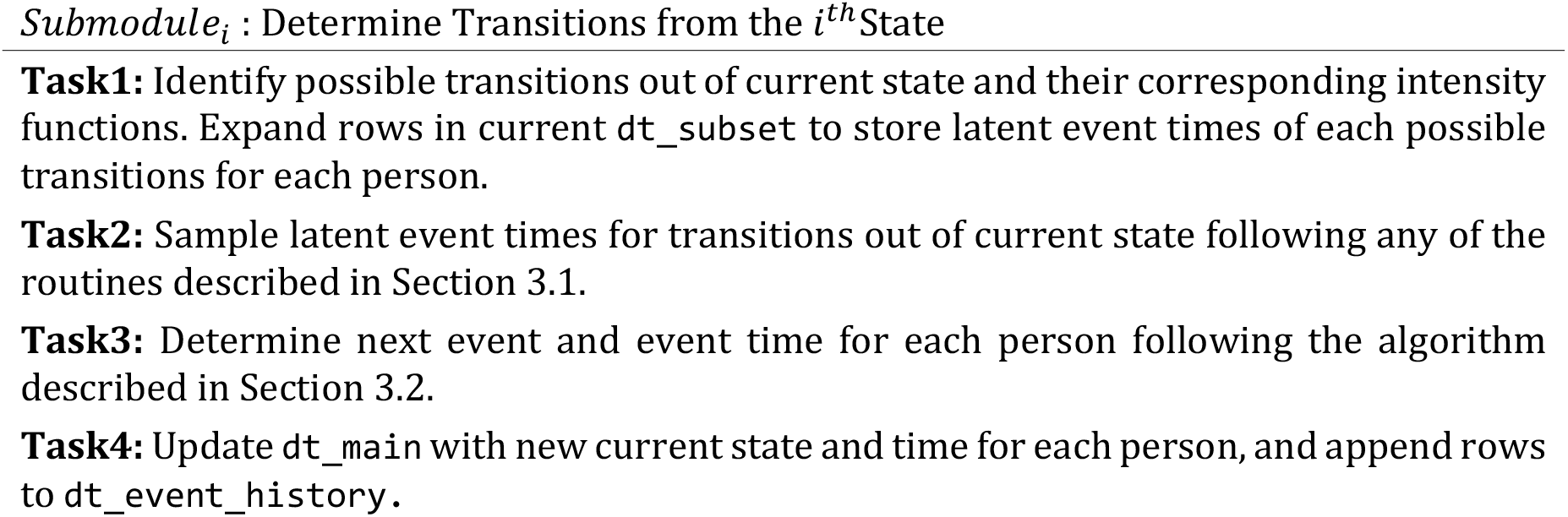
Structure and Tasks of Submodules in Module 2.

##### 5.3.2.1 Submodule 1: Transitions from Healthy

**Task 1:** Identify possible transitions from Healthy and expand dt_subset to include rows for the latent event times from H, creating dt_subset_long.

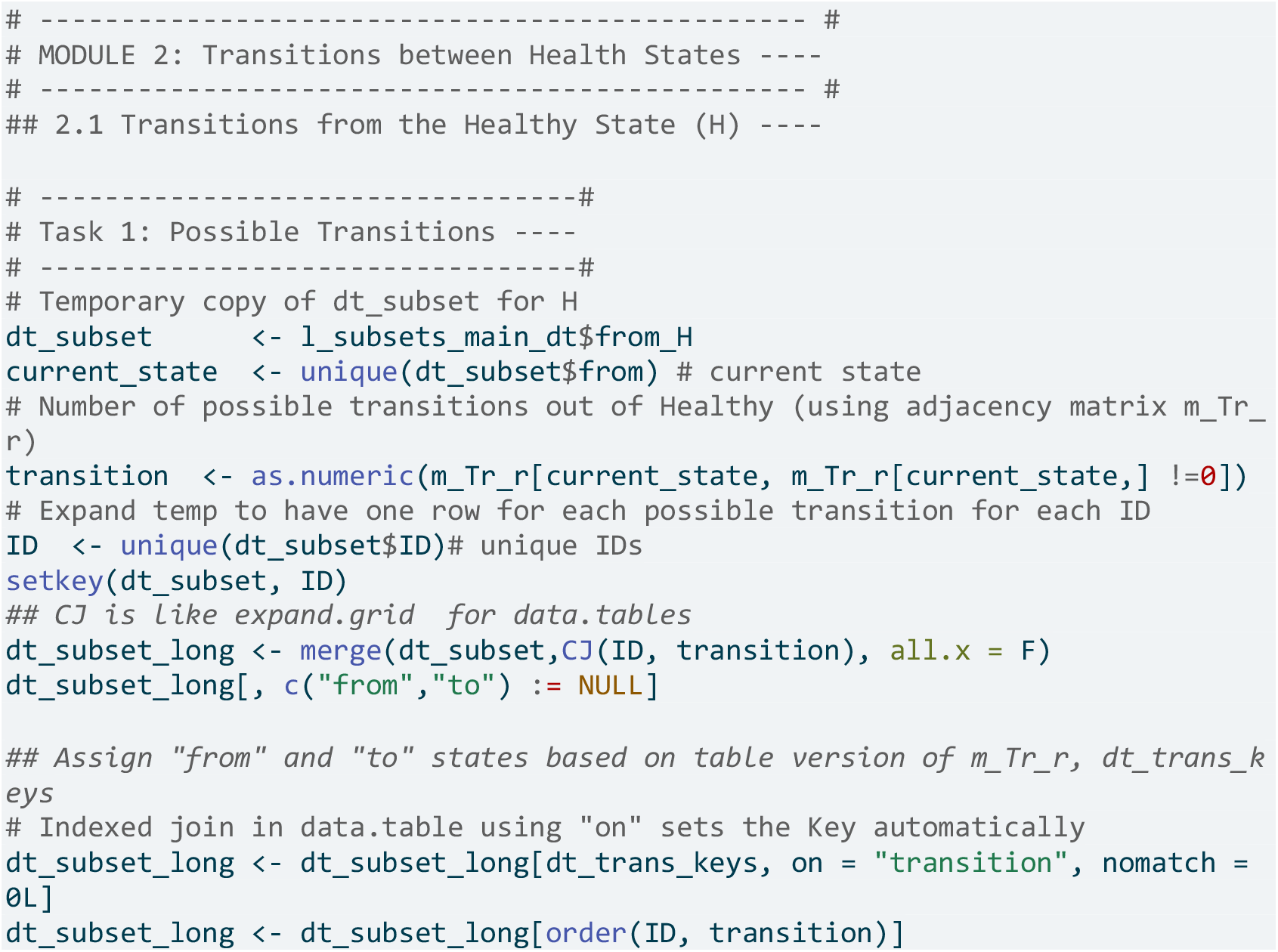

**Task 2:** Sample latent inter-event times (*τ*), for transitions out of Healthy, following two routines from Section 3.1.

- Subtask 2.1 H -> S : Healthy individuals face a constant annual rate of getting sick, implying that *τ*_*H,S*1_ *∼ exp*(*λ*_*H,S*1_). We use R’s base function ‘rexp()’ to sample this latent inter-event time *τ*_*H,S*1_, employing the inversion method.

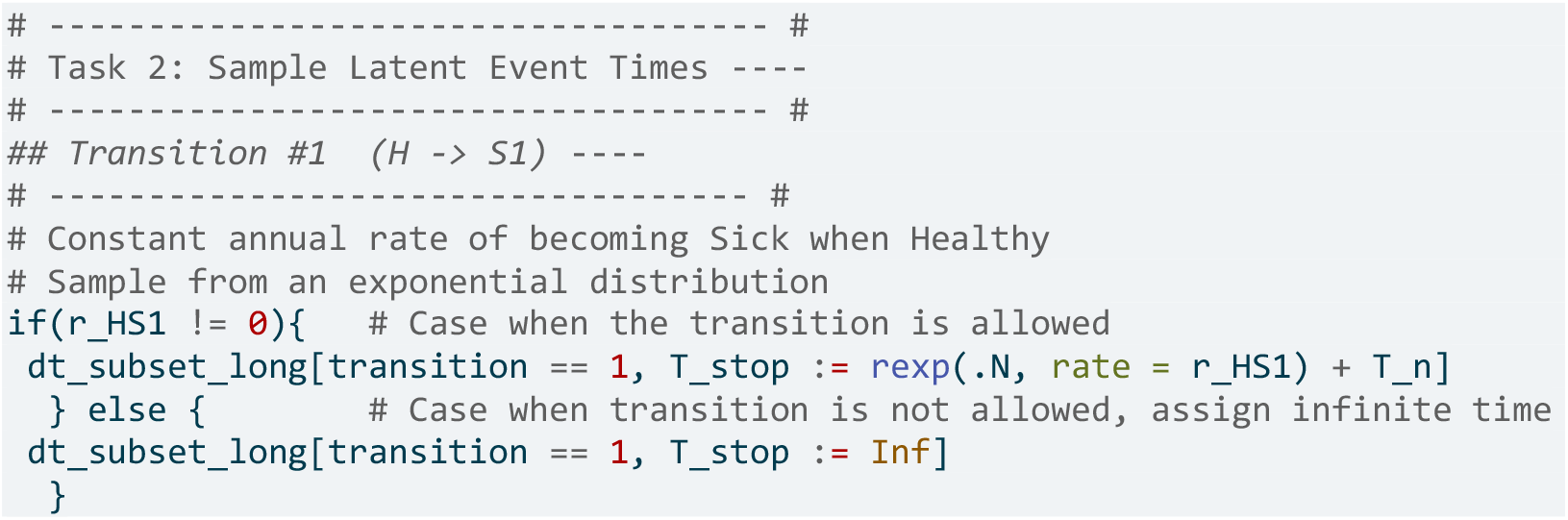

Subtask 2.2 H -> D : Healthy individuals face an age-specific background mortality. We sample latent times to death from dt_bckgrd_mortality using non-parametric sampling (Section 3.1).

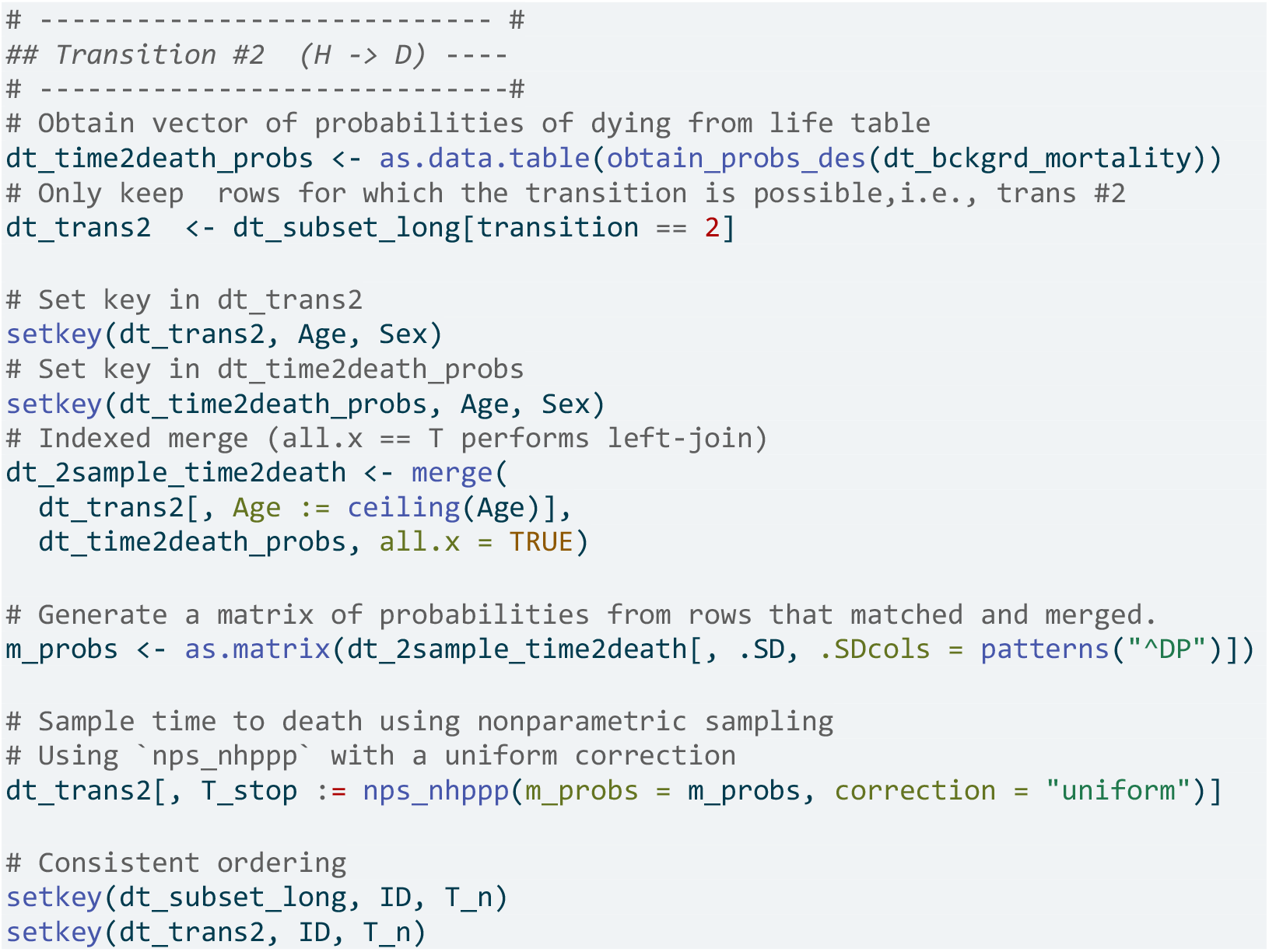
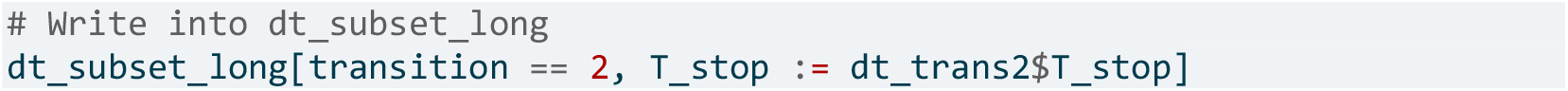

**Task 3:** Use samples of latent event times for all possible transitions per individual in the Healthy state to identify each individual’s next transition following the algorithm in Section 3.2.

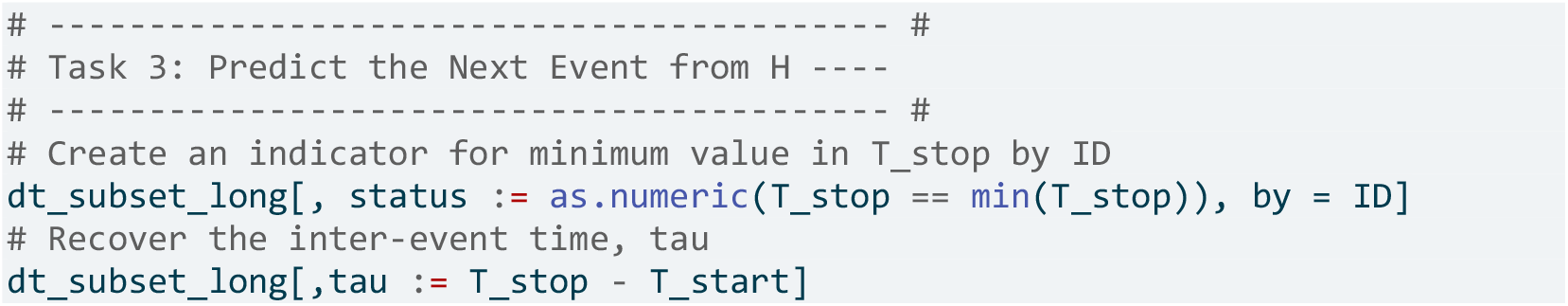

**Task 4:** Update each individual’s simulation time and age, which are needed for consistent sampling time to death. Add the predicted inter-event time from **Task 3** to the current simulation time.

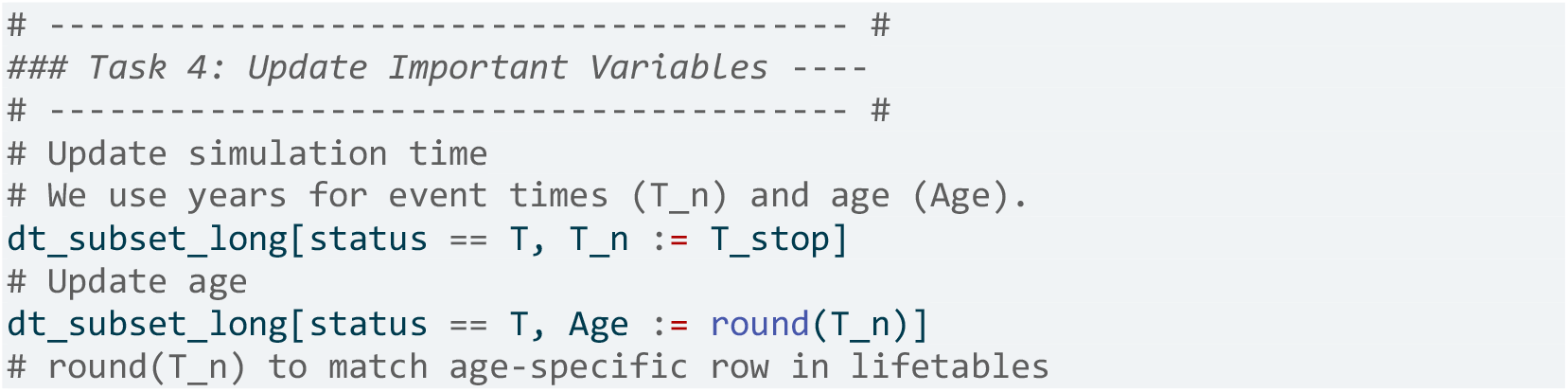

We track events histories, appending the new event information to dt_event_history in long format.

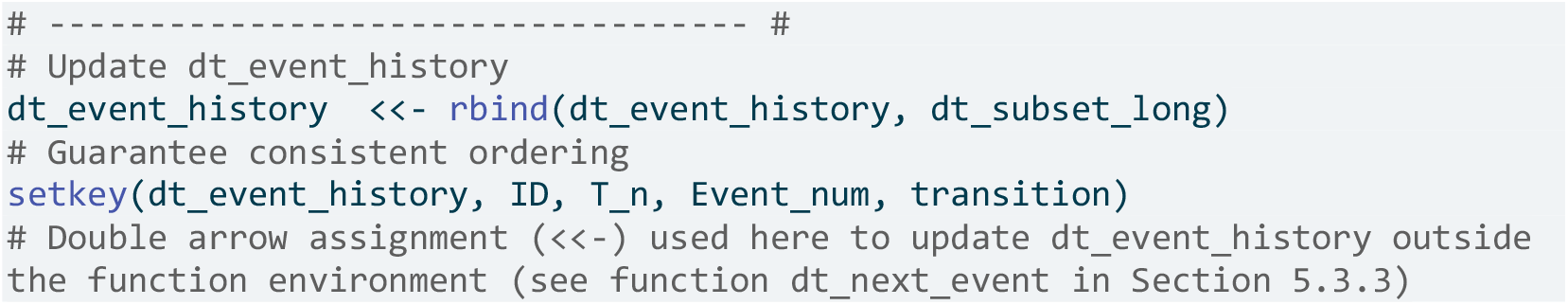

##### 5.3.2.2 Submodule 2: Transitions from Sick

Recall that Sick individuals’s hazard of progressing to the Sicker state increases as a function of time spent Sick. They also face a constant recovery rate to Healthy and increased age-dependent mortality while Sick.

We follow the same structure as in the submodule for Healthy but modify **Task 2** to sample the latent inter-event times from Sick to Sicker using a Weibull distribution with parameters `r_S1S2_scale` and `r_S1S2_shape`, latent inter-event times from Sick to Healthy from an exponential distribution with rate `r_S1H`, and time to death while Sick from the life table and hazard ratio. We omit the code for other Tasks, as they are mostly unchanged (see the appendix and GitHub repository).

**Task 2:** Sample latent inter-event times for transitions out of Sick following the two routines in Section 3.1. To include the effect of strategies “B” and “AB” on reducing progression to Sicker, we scale the baseline scale parameter of the Weibull distribution by `hr_S1S2_trtB`, assuming proportional hazards (Section 4.2).

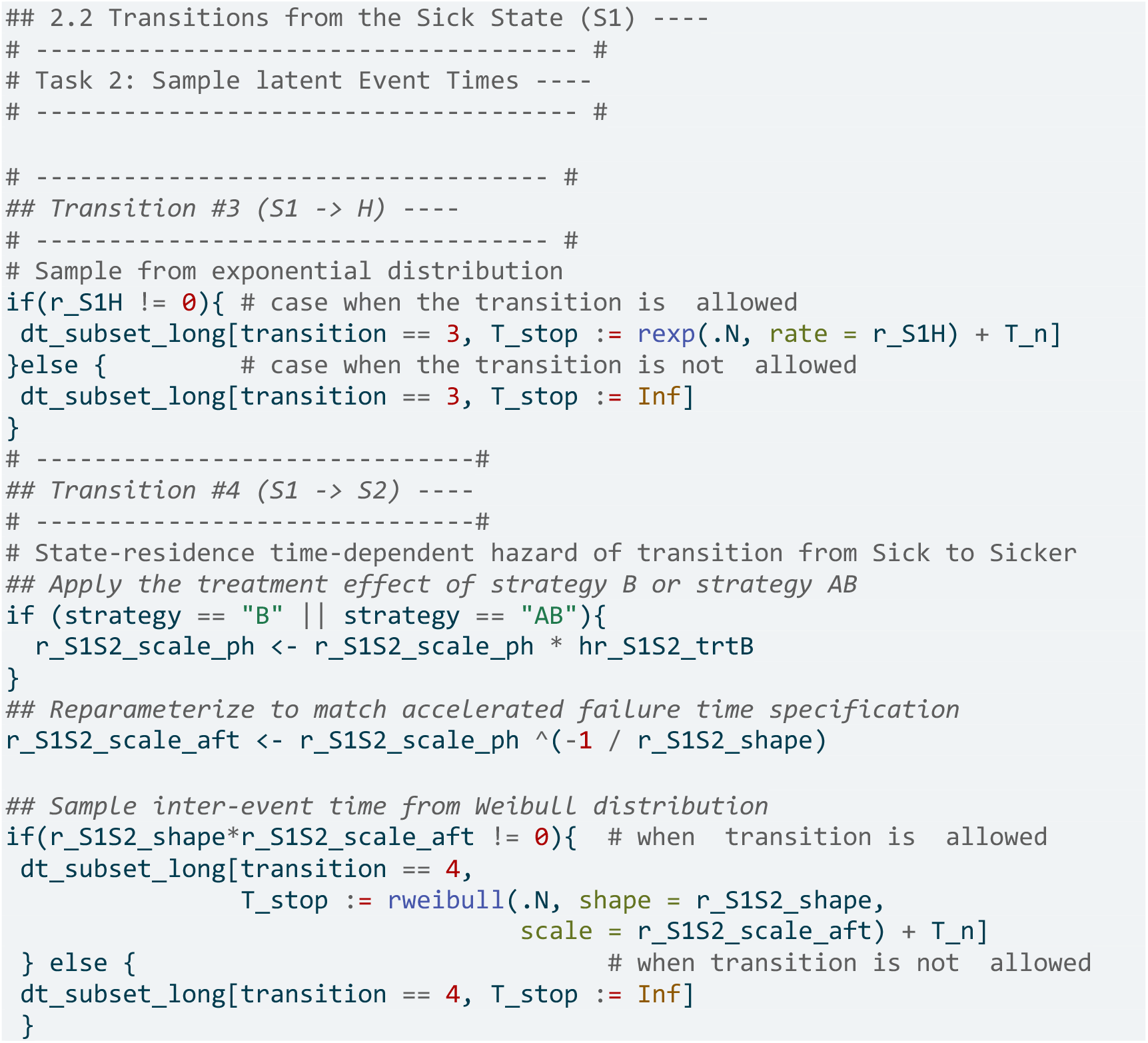

For sampling the time to death while Sick, we use the same non-parametric routine as in **Submodule 1**, but replace `dt_bckgrd_mortality` with the health-state-specific life table `dt_mortality_Sick` generated in **Module 1**.

##### 5.3.2.3 Submodule 3: Transitions from Sicker

This submodule follows the same structure as before but restricts the only transition from Sicker to Dead, so **Task 3** is unnecessary. Like **Submodule 2, Task 2** uses the life table for the Sicker state from **Module 1**, `dt_mortality_Sicker`.

#### 5.3.3 Module 3: Dealing with Recurrent Events

In a purely progressive STM without transitions from Sick to Healthy, the maximum number of iterations of Modules 1 and 2 are fixed, ensuring all simulated individuals die or reach 100 years. However, allowing a transition back to Healthy implies creating loops, making the maximum iterations of Modules 1 and 2 (now a random variable) unknown.

This module requires two components. First, wrap Modules 1 and 2 in `‘sim_next_event()’`. Here, we only show the structure, see the appendix and the GitHub repository for details.

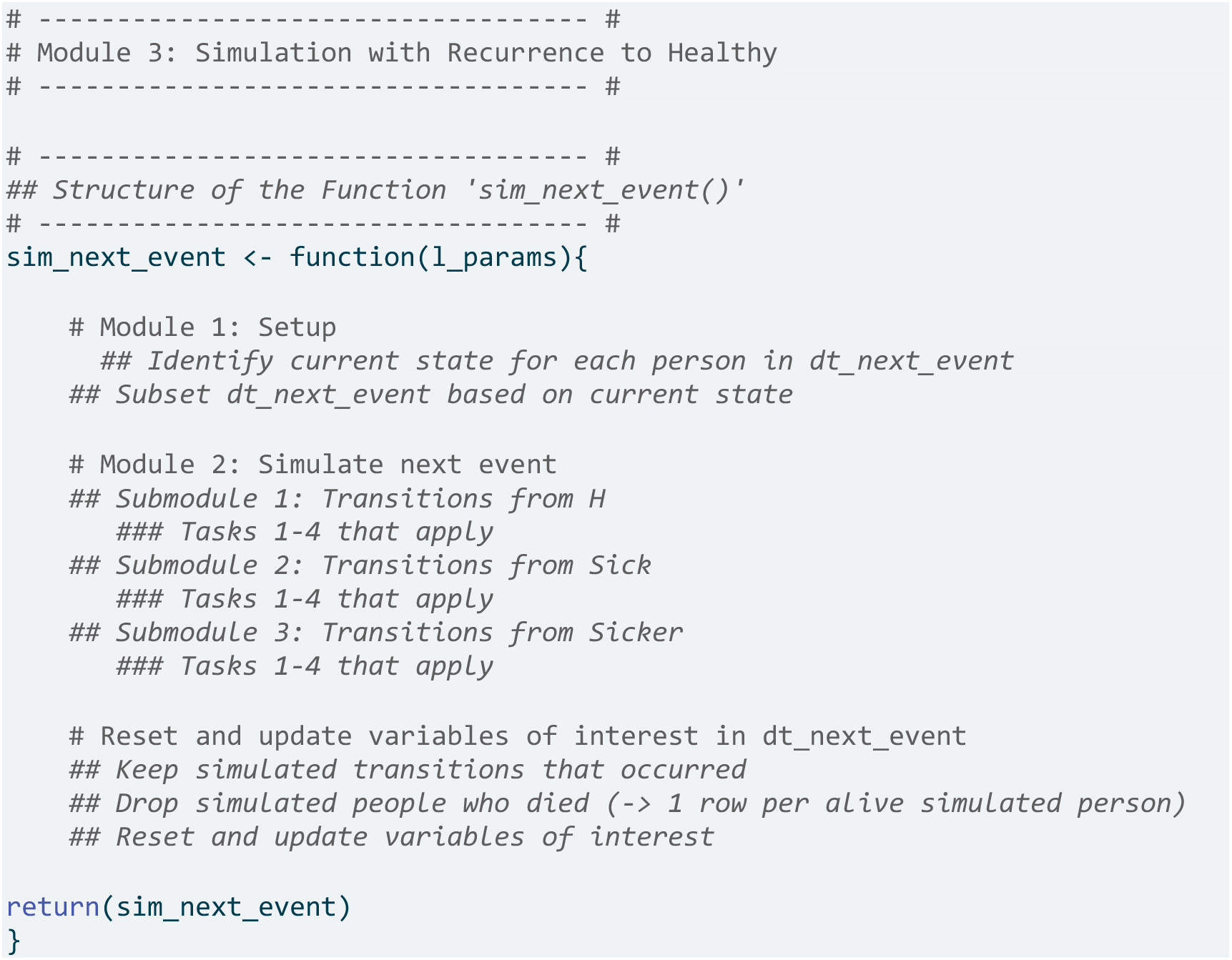

Second, we run a **while** loop that executes `sim_next_event` until everyone in the simulation has died or reached 100 years of age. We also set a **warning** for exceeding the maximum number of events as a sanity check.

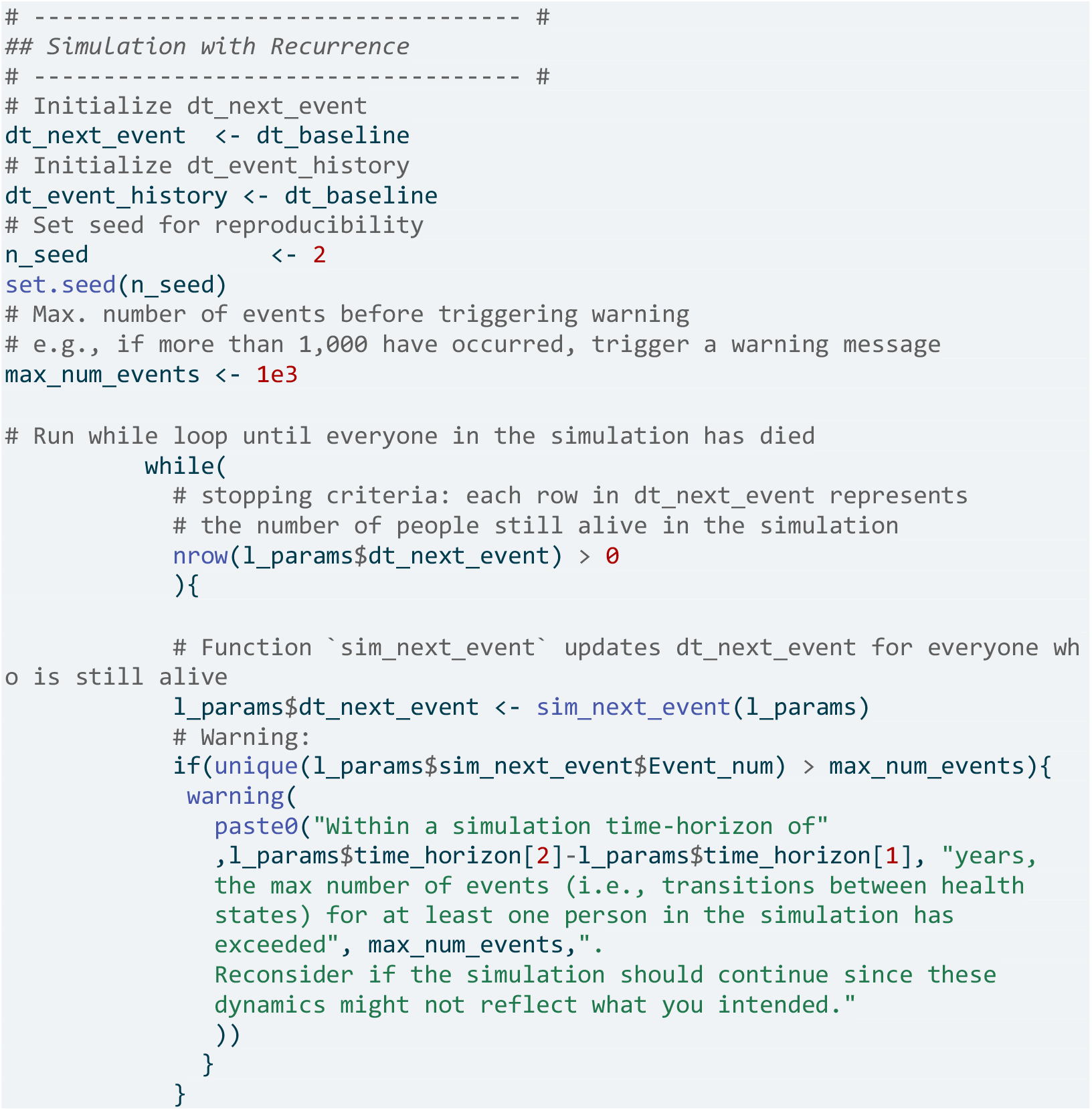

### 5.4 Post-processing and Auxiliary Modules

Below are code snippets illustrating the main components used to process the DES results and calculate outcomes. For the complete code to generate cost-effectiveness and epidemiological outcomes, refer to the appendix and GitHub repository.

#### 5.4.1 Post-processing Module A: Compute a Cost-effectiveness Analysis

This module computes a CEA of the four strategies. First, it generates the event history, `dt_event_history`, for each strategy using a **for** loop over `v_strategies`, executing **Module 3** each time.

We set a common random number (CRN) stream across all strategies to reduce variance in Monte Carlo estimates of incremental costs and benefits. [30] The event history tables are stored in `l_event_history_strategies`, which feeds into **Module A** for the CEA.

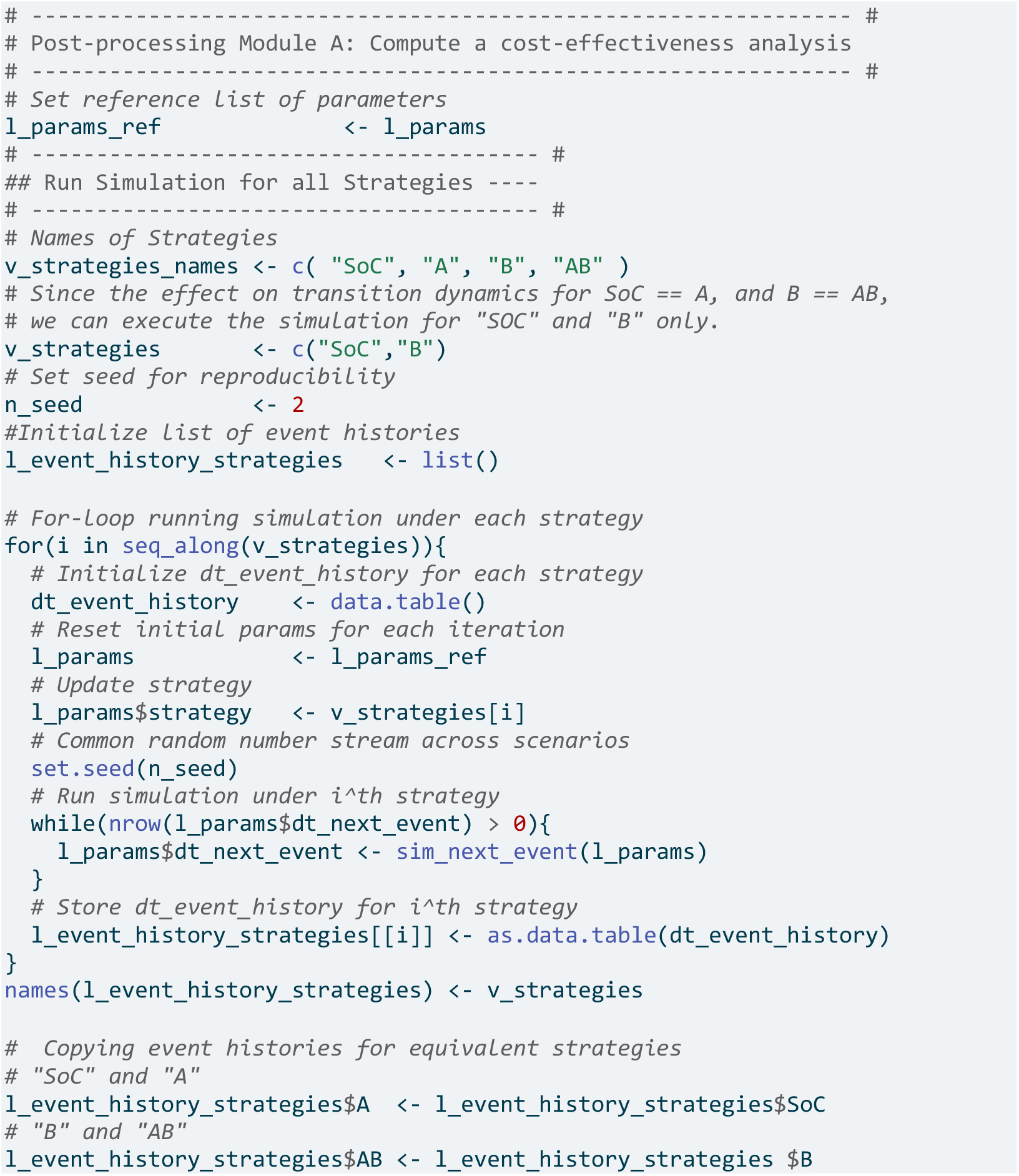

Next, we compute the expected discounted total costs and utilities under each strategy by appending each dt in `l_event_history_strategies` and creating `dt_all`, containing the four event histories. Costs and utilities are assigned to the time spent in each non-absorbing state, as well as for each treatment and transition between states (see Section 5).

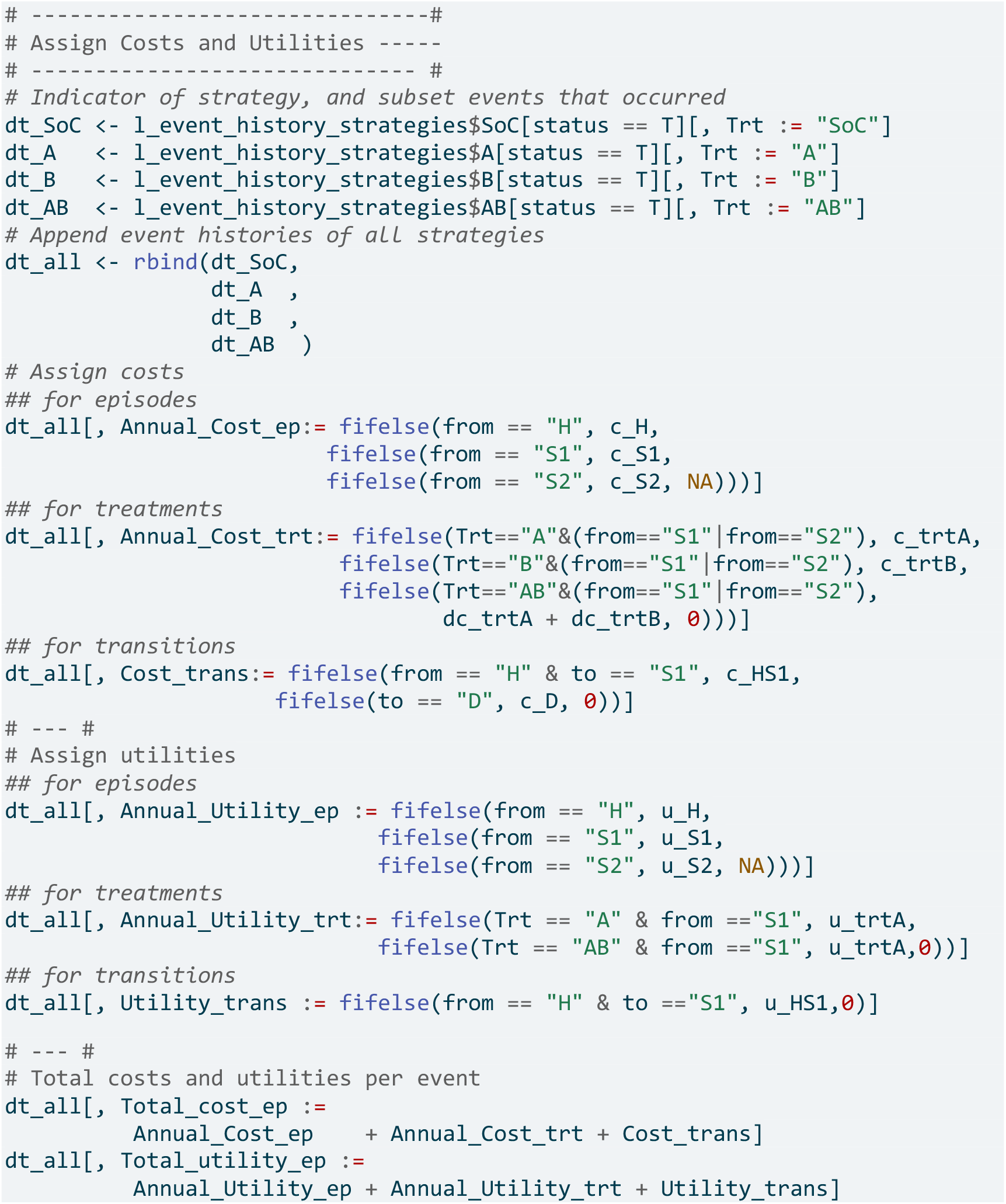

We exponentially discount all outcomes in continuous time. Let *O*_*i*_ be a cumulative outcome for the *i*^*th*^ individual for the interval [*t*_1_, *t*_2_], and *ρ* the exponential discount rate, then the discounted cumulative outcome over [*t*_1_, *t*_2_] for the *i*^*th*^ individual, 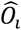, is:

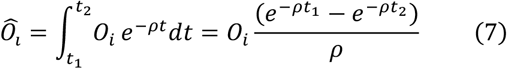

For discounting, the initial time is 0. We shift all inter-event times by subtracting the start age of the cohort (25 years of age), i.e., the time after birth when the simulation starts for each individual.

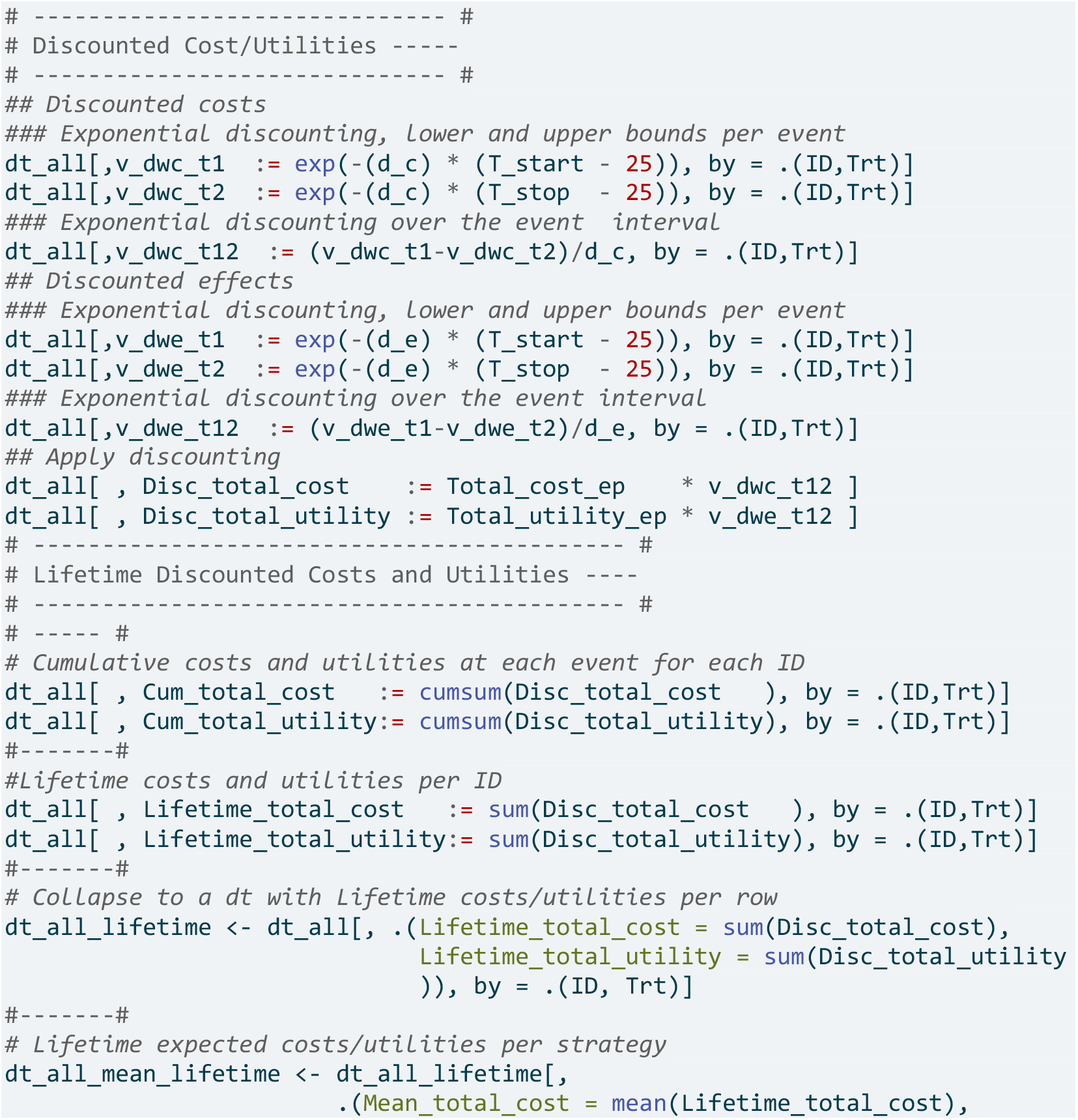
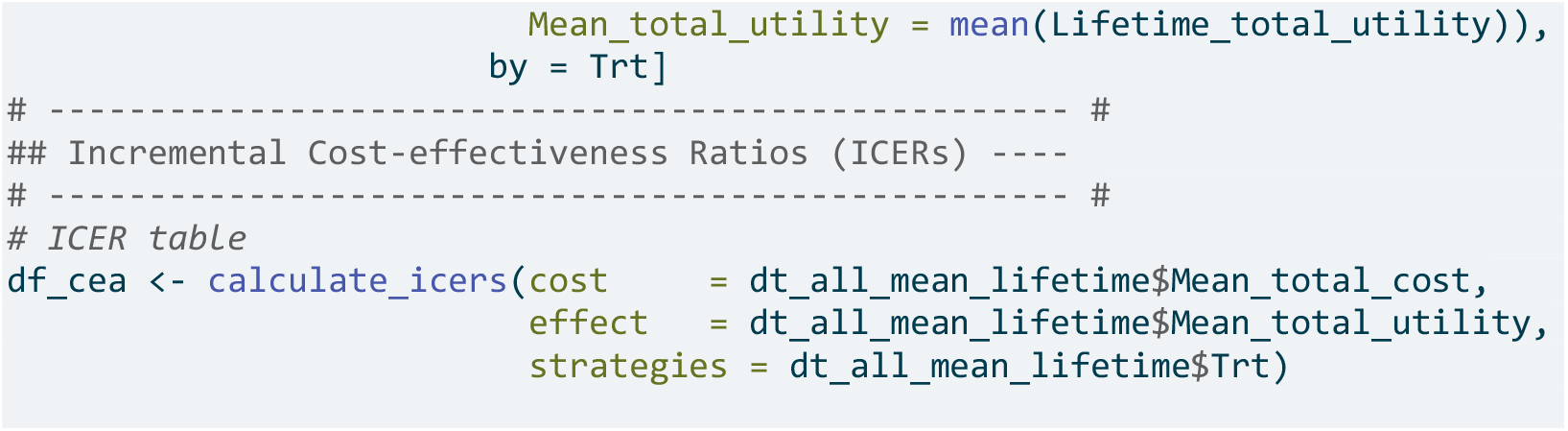

#### 5.4.2 Post-processing Module B : Compute Epidemiological Outcomes

Survival probabilities and disease prevalence are epidemiological outcomes obtainable directly from cSTMs and DES. Another outcome is dwell time in a specific health state, like time spent in the recurrence-free state in cancer models. Due to the time-to-event formulation of DES, dwell times at each alive state for each individual and for the cohort are directly obtained from the DES output. In this module, we compute survival probabilities, disease prevalence, and the distribution of dwell times for the entire cohort at each health state in the Sick-Sicker Model.

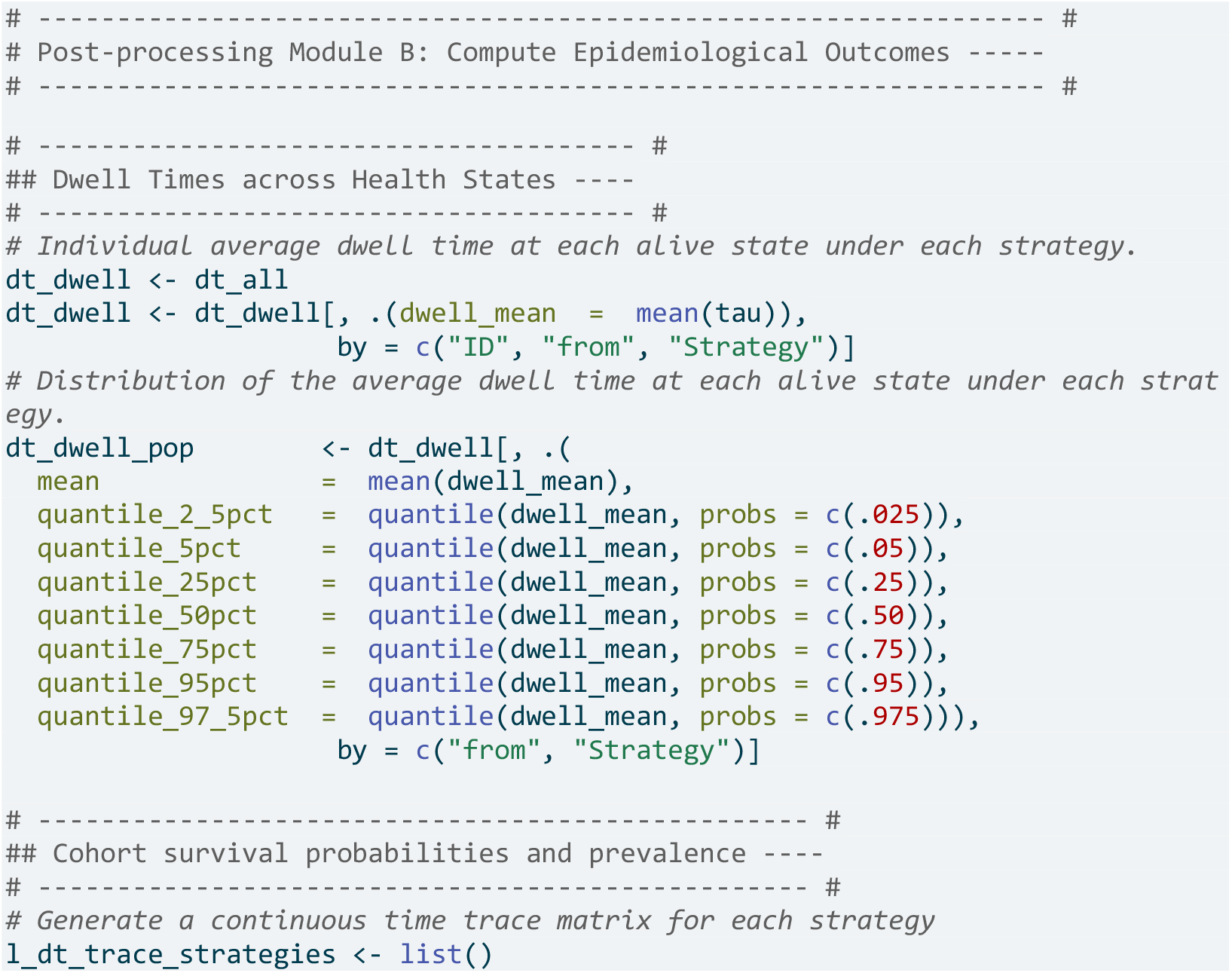
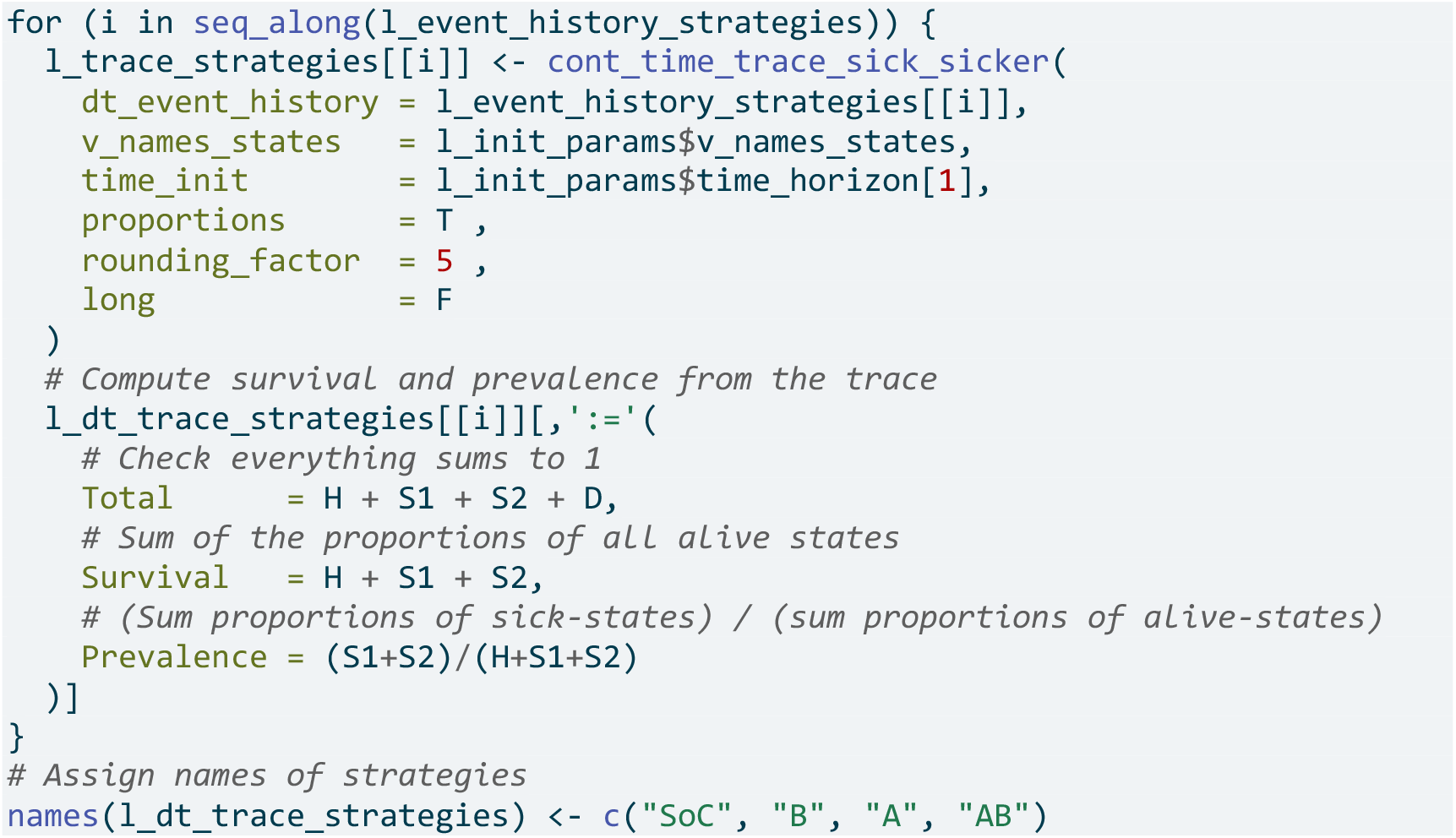

#### 5.4.3 Post-processing Module C: Compute a Probabilistic Analysis

This Module computes a probabilistic analysis (PA) to assess how model parameter uncertainty affects cost-effectiveness outcomes. It samples parameter sets using Latin Hypercube sampling[31] and user-specified distributions. Each set is used to simulate strategies and calculate their cost-effectiveness outcomes, stored in a PA data set consistent with previous tutorials [11, 12]. This compatibility allows use of validated functions for sampling parameter sets, and outcome calculations.

Figure 4 illustrates the CEA of the Sick-Sicker Model, with the scatter plot (See Panel A), cost-effectiveness acceptability curves (CEACs) and cost-effectiveness frontier (CEAF) (See Panel B), the expected loss curves (ELCs) (See Panel C), and the expected value of perfect information (EVPI). We sampled 1,000 parameter sets for PA results.

**Figure 4.**
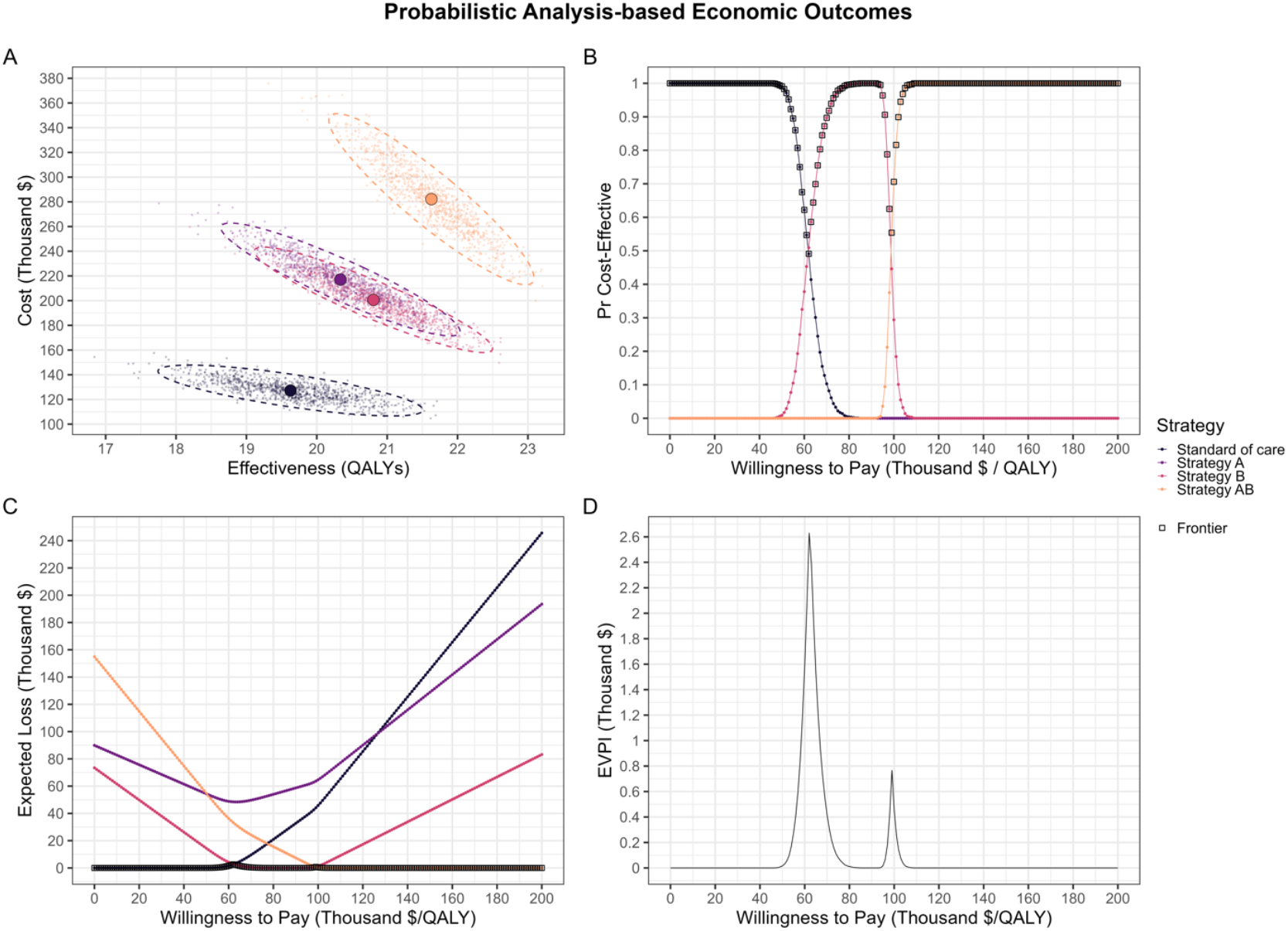
Probabilistic Analysis-based Economic Outcomes Panel A. Cost-effectiveness scatter plot shows pairs of expected costs and QALYs under each strategy for each parameter set. Panel B. Cost-effectiveness acceptability curves (CEACs) represent the probability that each strategy is cost-effective at each willingness to pay (WTP) threshold, and the cost-effectiveness frontier (CEAF), representing the probability of the optimal strategy being cost-effective at each WTP threshold ($1,000 intervals). Panel C. Expected loss curves (ELCs) represent the foregone benefits of choosing a suboptimal strategy at each WTP threshold. Panel D. Expected value of perfect information (EVPI) represent the maximum amount a decision-maker is willing to pay to obtain perfect information on all parameters of the decision model.

#### 5.4.4 Auxiliary Module: Continuous Time Trace Matrix and State Occupancy Plot

In the appendix, we include a module with functions that map the DES output to a cohort trace matrix (i.e., the proportions of simulated people in each state over time). This function enables quick calculations of epidemiological outcomes like survival probabilities and prevalence, similar to a discrete-time cSTM. Analysts can compare the proportions at each state over time from the DES with those simulated using discrete-time cSTMs. We also generate state occupancy plots to visually assess how the DES captures transition dynamics and the convergence of Monte Carlo estimators of state proportions over time.

### 5.5 Convergence of Monte Carlo Estimator

Finally, we explore how many individuals to simulate with the DES to produce estimates that converge to those obtained from an equivalent cSTM (mean-process model). In this example, we focus on DES-based estimators of the proportions of people in each state over time, along with their variability expressed as bootstrap confidence bounds at the 2.5^th and 97.5^th percentiles around the median. We showed that the variability of these state-occupancy estimates declines rapidly as the number of simulated individual by the DES increases (Figure 5). An implementation of this analysis in a distributed setting is available in the appendix and Github repository.

**Figure 5.**
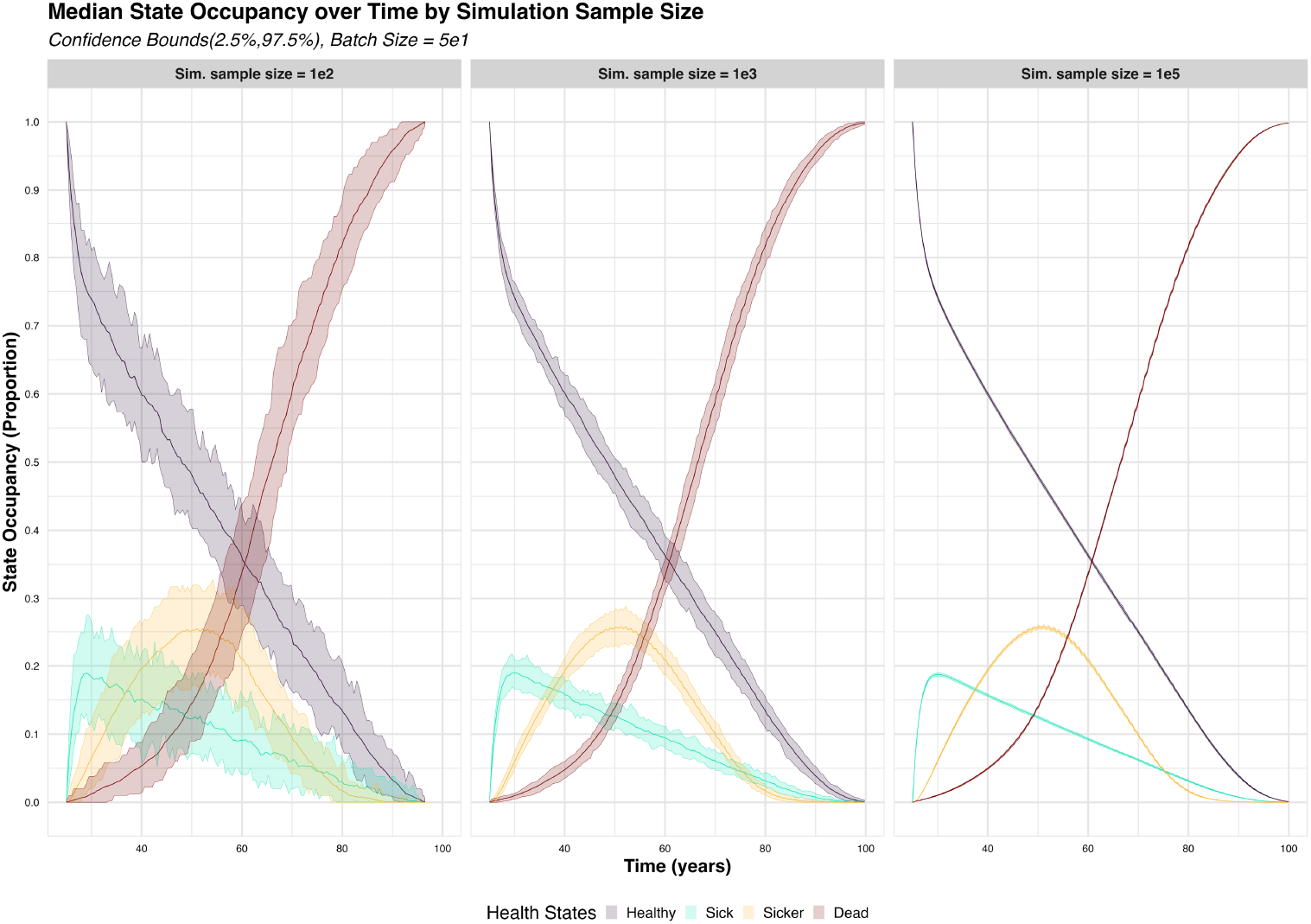
Median State Occupancy over Time by Simulation Sample Size State occupancy is calculated as the proportion of people in a given health state at a given time. We compute median, 2.5%, and 97.5% percentiles of state occupancy estimates by running fixed-sized batches of 5e1 independent runs of the simulation for cohorts of different sample sizes: 1e2, 1e3, 1e5. The variability of the Monte Carlo estimate of state occupancy decreases as we increase the simulation sample size.

## 6. Discussion

This tutorial introduces a novel, open-source DES framework designed for modeling realistic continuous-time, individual-level STMs. It supports features such as simulation and state-residence time dependence, treatment effects and other individual-level characteristics. We demonstrate the framework’s versatility and advantages within HTAs and medical decision making by providing a case example of a CEA in R using the previously published Sick-Sicker model.

DES offers computational advantages in terms of simulating models in which transition dynamics depend on state-residence time (i.e., the type of dynamics for which cSTMs expand their state space using “tunnel” states, see the tutorial paper by *Alarid-Escudero et al*., *2023)*. Alternative approaches to this for multi-state cohort models with semi-Markov dynamics include the use of Kolmogorov’s forward equations to calculate state-occupancy probabilities [32].Our individual-level DES samples event times from hazard functions, handling path dependence, competing risks, and time-varying covariates without solving integro-differential equations, providing a flexible alternative.

We demonstrate how treatment effects and individual-level characteristics influencing health-state transitions can be incorporated via a proportional-hazards approach, by scaling intensity functions. Incorporating heterogeneity in cSTMs adds computational load since the model assumes cohort homogeneity, requiring to to run as many simulations as the number of subpopulations. Other discrete-time methods like iSTMs may incorporate heterogeneity and individual-level factors without multiple runs, but DES offers a computational advantage by evaluating health states only at the times events occur. Our DES framework also captures within- and between-person variability in dwell times without complex calculations. For example, to assess personalized cancer treatments prolonging recurrence-free states, recovering dwell time distribution is crucial. cSTMs only provide mean dwell times, while discrete-time iSTMs recover both the population and individual variability. With DES, individual dwell times are directly obtained and can be summarized for the entire cohort in a few code lines.

When implementing a DES, there are important analytic choices. Intensity functions must accurately reflect the joint distribution of time-to-event for each transition, which can be achieved by tracking individuals’ times and states. A multistate model can then estimate transition intensities considering competing events [16, 33]. Using estimates from different studies and populations can misrepresent dynamics, potentially biasing estimates of epidemiological and economic outcomes. Calibration is necessary to match key epidemiological outcomes of the target population. Additionally, the probabilistic analysis should quantify how uncertainty in the transition rates, e.g., uncertainty on the hazard ratios that affect mortality, affects decision making.

Deciding whether to use specialized software (like ‘simmer’ [34, 35], ‘quecomputer’ [36], ‘microsimualtion’ [37] and ‘hesim’ [38]) or lower-level code to implement DES is important. Our DES implementation offers a transparent, domain-specific reference for individual-level, continuous-time state-transition modeling in health economics, focusing on accessibility, auditability, modifiability, and integration with survival analysis and probabilistic analysis workflows in R. It complements general DES packages suited for resource/queue management or high performance. This tutorial uses a specific coding and naming convention for readability, consistency, and ease of extension, but choosing any suitable convention is acceptable and should align with team practices.

iSTMs, whether simulated in discrete or continuous time like DES, may bias estimates of decision uncertainty and value of information. Goldhaber-Fiebert et al 2025 provide a closed-form and numerical characterization of this bias, showing the bias is asymptotically consistent and can be minimized by increasing the number of simulated individuals [39]. Analysts using DES should assess the number of simulated individuals needed to reduce bias to tolerably low levels. To keep the tutorial focused, we used a single random number seed generating a common random number (CRN) stream across strategies as a variance-reduction technique, requiring smaller samples [30]. Analysts conducting comparative experiments may further improve precision by mapping random number sub-streams (RNGS) (e.g., MRG32k3a) to distinct processes and synchronizing RNGS across scenarios, but requires careful design to avoid unintended correlations and maintain event-level alignment across scenarios.

This tutorial provides an open-source and transparent framework for stochastic simulation of individual, state-transition models in continuous time via an event-driven approach. It explains the key mathematical concepts and provides a detailed guide to perform a CEA from first principles, generate epidemiological outcomes, and conduct a probabilistic analysis, including EVPI, required by CEA panels and multiple HTA agencies. Decision scientists using modeling approaches like DES can benefit from understanding the mathematical concepts, coding techniques, and design that our framework illustrates, regardless of whether they use specialized software. The concepts and techniques in DES are closely related to those in simulating cSTMs and iSTMs in discrete time, with significant gains in computational efficiency and flexibility in modeling realistic dynamics.

## Supporting information

Appendix_Rcode

## Data Availability

Synthetic data and code are available online at https://github.com/LopezM-Mauricio/DES_tutorial_timedep_SickSicker

