## Appendix_supplemental_materials for "A Tutorial on Discrete Event Simulation Models in R using a Cost-Effectiveness Analysis Example"

**eTable 1: Execution time Distribution for the Recursive Sick-Sicker Model by Simulation Sample Size.**

| **Execution Time Distribution (minutes)**<br>(50 replications for each sample size) | | | | | |
|---|---|---|---|---|---|
| **Simulation Sample Size (in thousands)** | 0.1 | 1 | 10 | 100 | 1,000 |
| **Minimum** | 0.005167 | 0.009833 | 0.05633 | 0.4968 | 4.797 |
| **0.25 Quantile** | 0.006333 | 0.014500 | 0.07083 | 0.5911 | 5.750 |
| **Median** | 0.008500 | 0.019500 | 0.08075 | 0.6974 | 6.566 |
| **Mean** | 0.012077 | 0.025753 | 0.09620 | 0.7366 | 6.862 |
| **0.75 Quantile** | 0.010292 | 0.021875 | 0.09379 | 0.7896 | 7.501 |
| **Maximum** | 0.197667 | 0.308333 | 0.49450 | 1.9805 | 14.884 |

**Benchmarking Details**

**CPU model:**
Intel® Xeon® CPU E5-2630 0 @ 2.30 GHz (2 processors)
**Memory Size:**
128 GB (RAM)
**Operating System:**
Microsoft Windows Server 2019
**R version:**
64-bit R-4.22
**R-Studio version:**
RStudio 2023.03.0+386 "Cherry Blossom" Release (3c53477afb13ab959aeb5b34df1f10c237b256c3, 2023-03-09) for Windows
Mozilla/5.0 (Windows NT 10.0; Win64; x64) AppleWebKit/537.36 (KHTML, like Gecko) RStudio/2023.03.0+386 Chrome/108.0.5359.179 Electron/22.0.3 Safari/537.36
**Benchmarking Tools:**
Base R function 'system.time()'
**Number of threads:**
21 (multi-thread data.tables)

### eTable 2: General and Specialized Dependencies by Module of the Coded Example

| Modules | Script Name in Repository | General Data Manipulation Dependencies | Specialized Simulation and Plotting Dependencies |
|---|---|---|---|
| **Lightweight Modules** | | | |
| 1. DES progressive | 'DES_Sick_Sicker_progressive.R' | 'data.table' ; 'dplyr' | Not applicable |
| 2. DES with recursion to the Healthy State | 'DES_Sick_Sicker_recurrence.R' | | Not applicable |
| 3. Epidemiological Outcomes | 'Module_B_Epi_Outcomes.R' | 'data.table' ; 'dplyr' | Not applicable |
| 4. Economic Outcomes | 'Module_A_CEA.R' | 'data.table' ; 'dplyr' | Not applicable |
| 5. State Occupancy over Time | 'Aux_Module_Cont_Time_Trace.R' | 'data.table' ; 'dplyr' | Not applicable |
| **Parallelized Modules** | | | |
| 6. Probabilistic Analysis | 'Module_C_Probabilistic_Analysis.R' | 'data.table' ; 'dplyr' | 'doParallel' 'parallel' 'foreach' 'MethylCapSig' 'abind' |
| 7. Simulation Sample Size Analysis | 'Aux_Module_Convergence.R' | 'data.table' ; 'dplyr' | 'doParallel' 'parallel' 'foreach' 'abind' |
| **Modules to generate Plots** | | | |
| 8. Economic Outcomes | 'Module_CEA_wPlots.R' | 'data.table' ; 'dplyr' | 'ggplot2' 'ggrepel' |
| 9. Epidemiological Outcomes | 'Module_Epi_Outcomes_wPlots.R' | 'data.table' ; 'dplyr' | 'ggplot2' 'patchwork' 'viridis' |
| 10. State Occupancy | 'Aux_Module_Cont_Time_Trace_wPlots.R' | 'data.table' ; 'dplyr' | 'ggplot2' 'viridis' |
| 11. Probabilistic Analysis | 'Module_Probabilistic_Analysis_wPlots.R' | 'data.table' ; 'dplyr' | 'doParallel' 'parallel' 'foreach' 'MethylCapSig' 'ggplot2' 'patchwork' 'viridis' 'ellipse' 'abind' |
| 12. Simulation Sample Size Analysis | 'Aux_Module_Convergence_wPlots.R' | 'data.table' ; 'dplyr' | 'doParallel' 'parallel' 'foreach' 'MethylCapSig' 'ggplot2' 'viridis' 'patchwork' 'abind' |

### eFigure 1 Structure and Logic of Coded Example

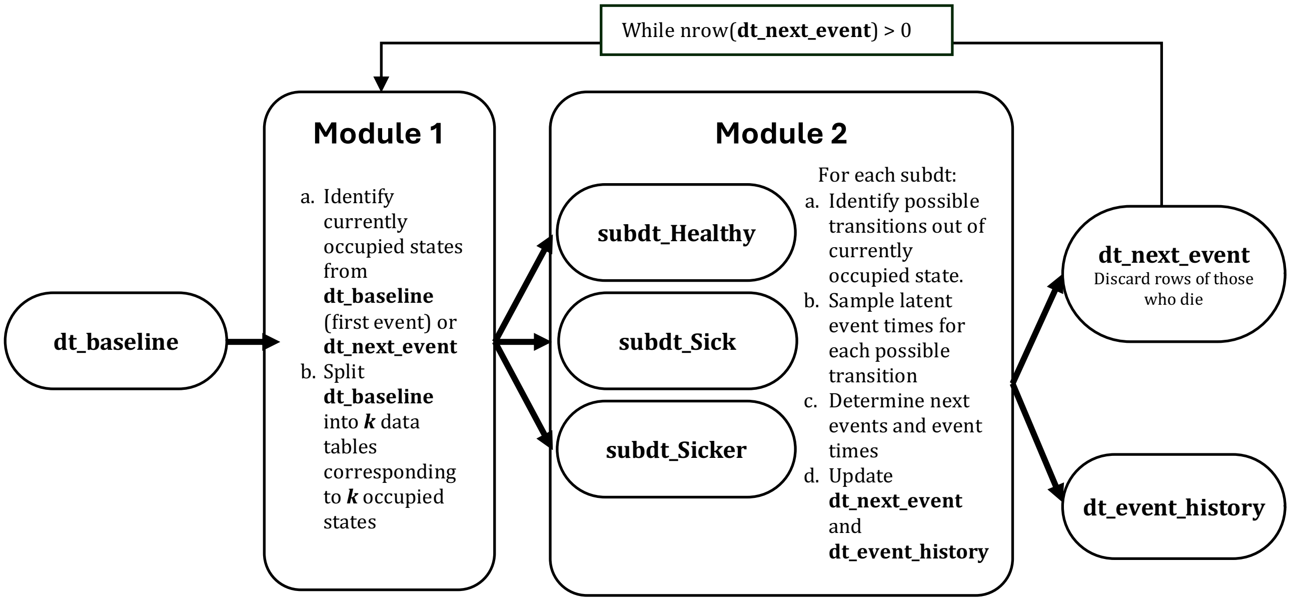


Steps for 1st Event:

a. **Module 1** takes **dt_baseline** as input, identifies the currently occupied health states (in this example, everyone starts at Healthy) and splits **dt_baseline** into **k** different subtables, one for each occupied state.
b. **Module 2** takes the **k** subtables as input (for the 1st event only **subdt_Healthy** is required) and for each subtable the module executes the tasks detailed in Table 2 of the main text to determine the next events and the times of the next events.
c. **Module 2** updates the state variables and time of events in datatables **dt_next_event** and **dt_event_history**. People who die are discarded from **dt_next_event** after this update.

Steps after the 1st Event:

a. **Module 1** takes **dt_next_event** as input, identifies the currently occupied states (now different states can be occupied) and splits **dt_baseline** into **k** different subtables, one for each occupied state.
b. **Module 2** takes the **k** subtables as input (after the 1st event any of **subdt_Healthy, subdt_Sick, subdt_Sicker** can be required) and for each subtable the module executes the tasks detailed in Table 2 of the main text to determine the next events and the times of the next events.

Stopping Criteria:

a. A 'while' loop feeds each new version of **dt_next_event** to **Module 1** until everyone has died, which occurs when **dt_next_event** is an empty table. Other stopping criteria are possible by modifying the conditions inside the 'while' loop.

Adapting the Code:

a. Initial occupied states can be arbitrarily set by modifying the corresponding column in **dt_baseline**.
b. Increasing the state space will increase the number of subtables generated as inputs for **Module 2,** which is done automatically, but requires manually specifying in **Module 2** the transition intensities of the new added states in the corresponding submodules for sampling latent event times.
c. Modifying transition dynamics, e.g., using another event time distribution for a specific transition, can be done by modifying submodules of **Module 2** that correspond to sampling of latent event times for each transition.
d. Covariates that inform transition intensities need to be included as columns in **dt_baseline** and carried over to **dt_next_event.**

### Data.table and other R Package Dependencies

In this tutorial, we use data.table (dt), an R object optimized for memory efficiency and computational speed for handling large data sets with millions of rows, and offers faster grouping, summarizing, and aggregation operations with fewer memory requirements than data.frame and tibble for the case example[1]. For an introduction to data.table and its syntax we refer the reader to open-source resources (e.g., see online [2] )Readers interested in the execution time distribution of the coded example for different simulation sample sizes can consult eTable 1 in this appendix. In addition to data.table, running the coded example only requires the 'dplyr' package, a well-maintained data-manipulation package in R, [3] making the implementation of the coded example lightweight.

To run the probabilistic analysis and the analysis of the required simulation sample size, it is suggested to run the parallelized version of the code that are provided in the GitHub repository. Distributed runs will require installing other packages, such as 'doparallel'. Similarly, reproducing the exact plots presented in the tutorial will require specialized packages. Readers can consult a detailed list of the dependencies required to run each Module of the coded example in eTable 2 in this appendix and in the GitHub repository.


[1] T. Barrett *et al.*, "data.table: Extension of `data.frame`." p. 1.17.0, Apr. 15, 2006. doi: 10.32614/CRAN.package.data.table.

[2] "Introduction to data.table." CRAN. Accessed: Apr. 02, 2025. [Online]. Available: https://cran.r-project.org/web/packages/data.table/vignettes/datatable-intro.html

[3] H. Wickham, R. Francois, L. Herny, K. Müller, and D. Vaughan, *dplyr: A Grammar of Data Manipulation*. (2025). [Online]. Available: https://CRAN.R-project.org/package=dplyr
