## Appendix_Rcode for "A Tutorial on Discrete Event Simulation Models in R using a Cost-Effectiveness Analysis Example"

---

#### 1. DES Implementation of a Progressive Sick Sicker Model

In this section we detail the code to implement a discrete event simulation of a purely progressive version of the Sick Sicker model. This section is based on Subsections `Module 1` and `Module 2` in the main text.

Complexity included in the model:

- Time Dependence: the DES incorporate two types of type dependency

---

[1] Department of Health Policy, Stanford School of Medicine, Stanford University, Stanford, CA, USA.

[2] Department of Health Policy, Stanford School of Medicine, Center for Health Policy, Freeman-Spogli Institute for International Studies, Stanford University, Stanford, CA, USA

- A. Simulation time-dependence (Age specific mortality)
  - B. State residency time-dependence (Transition S1 -> S2)
- Treatment effect acting on transition rate S1 -> S2

```
#------------------------------------------------------------------------------
--#
# Initial setup ----
rm(list = ls())    # remove any variables in R's memory
gc()               # clean working memory
#------------------------------------------------------------------------------
--#
## Load packages ----
library("data.table"  )
library("dplyr"       )

## Load DES-specific and utility functions ----
source("R/DES_functions.R") # DES specific
source("R/Functions.R")     # General utility functions

#------------------------------------------------------------------------------
--#
#* To program this tutorial we used:
#* R version 4.2.1 (2022-06-23)
#* Platform: aarch64-apple-darwin20 (64-bit)
#* Running under: Mac OS 12.5
#* RStudio: Version 2022.07.1+554
#------------------------------------------------------------------------------
--#
# Description ----
#* This code implements a Discrete Event Simulation (DES) of a purely progres
sive
#* Sick-Sicker model.
#* Structure: Code is structured in modules as described in Lopez-Mendez et a
l 2025.
#* Complexity included in the model:
#* Time Dependence: the DES incorporate two types of type dependency
#* A. Simulation time-dependence (Age specific mortality)
#* B. State residency time-dependence (Transition S1 -> S2)
#*Treatment effect on transition  S1->S2
#------------------------------------------------------------------------------
--#
# Initial setup ----
rm(list = ls())    # remove any variables in R's memory
gc()               # clean working memory
#------------------------------------------------------------------------------
--#
## Load packages ----
library("data.table"  )
library("dplyr"       )
```

```
## Load DES-specific and utility functions ----
source("R/DES_functions.R") # DES specific
source("R/Functions.R")     # General utility functions

#-----------------------------------------------------------------------------
--#
# Initialize Parameters ----

# Simulation Parameters
## Simulation Time horizon
time_init        <-   25
time_end         <-   100
time_horizon     <- c(time_init,time_end)
# Simulation sample size
n_sim            <-   1e3                          # Simulation sample si
ze

## Model State Space and Transitions
v_states         <- c("H",                         # Healthy (H)
                      "S1",                        # Sick (S1)
                      "S2",                        # Sicker (S2)
                      "D")                         # Dead (D)
## Indexed Adjacency Matrix
# Zero entries indicate no transition between pairs of states,
# Nonzero positive entries index possible transitions in arbitrary order
m_Tr_r           <- matrix(c(0,1,0,2,
                             3,0,4,5,
                             0,0,0,6,
                             0,0,0,0),
                           nrow = 4, ncol = 4, byrow = TRUE,
                           dimnames = list(c("H", "S1","S2","D"),
                                           c("H", "S1","S2","D")))
## Indexed transitions, tabular
dt_trans_keys    <- data.table()
dt_Tr_r          <-  as.data.table(m_Tr_r)
dt_Tr_r[, from    := v_states]
dt_trans_keys    <- as.data.table(melt(dt_Tr_r, id.vars = "from",
                                       measure.vars = c("H" ,"S1" ,"S2" ,"D")
,
                                       variable.name = "to",
                                       value.name = "transition"))
dt_trans_keys    <- dt_trans_keys[transition !=0,][order(transition)]

## Transition Rates

### Mortality, specify subfolder in project to load Life Tables
lifetable            <- "Inputs/all_cause_mortality.Rda"
load(lifetable)
dt_bckgrd_mortality   <- as.data.table(all_cause_mortality)
```

```
dt_bckgrd_mortality   <- dt_bckgrd_mortality[,death_rate :=  Rate][Year== 201
5]
### Mortality Hazard Ratio while being S1
hr_S1                 <-    3
### Mortality Hazard Ratio while being S2
hr_S2                 <-    10

### Other transitions
### H  -> S1, constant rate ~Exp(r_HS1)
r_HS1                 <-    0.15
### S1 -> H,  constant rate ~Exp(r_S1H)
r_S1H                 <-    0.5
### S1 -> S2, state-residency time dependent transition
# tau ~ Weibull(r_S1S2_scale_ph, r_S1S2_shape)
# PH parameterization
r_S1S2_scale_ph       <-    0.08
r_S1S2_shape          <-    1.1
# Strategy
# Can be any of "SoC", "A", "B", "AB". Only B and AB, affect transitions rate
s
strategy              <-    "SoC"
## Effectiveness of strategy B and strategy AB
# hazard ratio of becoming Sicker when Sick under treatment B or AB
hr_S1S2_trtB          <- 0.6

#-----------------------------------------------------------------------------
--#
#-----------------------------------------------------------------------------
--#
# Useful Data tables
# Initialize `dt_baseline`
age_init         <- 25                         # Initial age of the cohort
## Sex group mortality rate
Sex_grp          <- "Total"
## Initial state
state_init       <- "H"
# Init dt
dt_baseline               <- data.table(
  # Covariate info
  ID                    = 1:n_sim,             # Numerical identifiers
  Age                   = age_init,            # Age at start
  Sex                   = Sex_grp,             # Sex, only if specified
  # Event tracking
  from                  = state_init,          # Origin state, first row eve
ryone starts in H, update accordingly at each event.
  to                    = NA,                  # Destination state, first ro
w everyone is NA, update accordingly at each event.
  T_n                   = time_horizon[1],     # Simulation time, records ti
me of events, first row all entries are zero, but will be updated to the time
when transition/event  occurs
```

```
  T_start              = time_horizon[1],        # State-residence time start, first row all entries are zero, but will be updated to the transition that occurs next
  T_stop               = 0,                      # State-residence time stop, first row all entries are zero, but will be updated to the transition that occurs next
  tau                  = 0,                      # Time-increment, first row all entries are zero, updated after each event
  status               = NA,                     # Occurrence/censoring status, transition-specific indicator, updated after each event.
  Event_num            = 1                       # Event number, at least 1 event from H-> D over time-horizon, updated after each event.
)

#----------------------------------------------------------------------------#
# Simulation (purely progressive) ----
# Initialize dt_next_event
dt_next_event      <- data.table()
# Initialize long-format dt_event_history
dt_event_history   <- data.table()
# Set seed for reproducibility
n_seed             <- 2
set.seed(n_seed)

# ---- #
#  Generate state-specific life-tables
## Inputs:   dt_bckgrd_mortality , and  hr_S1, hr_S2
## 2.1 Create l copies of the original life table, l corresponds to the number of states with a transition to dead
## 2.2 Apply state-specific hazard ratios to the original life table to obtain state-specific life tables.
## Mortality life tables
### Mortality from Sick
dt_mortality_Sick        <- copy(dt_bckgrd_mortality)
dt_mortality_Sick[, death_rate := death_rate*hr_S1]
### Mortality from Sicker
dt_mortality_Sicker      <- copy(dt_bckgrd_mortality)
dt_mortality_Sicker[, death_rate := death_rate*hr_S2]

#----------------------------------------------------------------------------#
# Transitions between Health States
#----------------------------------------------------------------------------#
# General Structure of the purely progressive Sick-Sicker Model:
#   |-> Module  1: Transitions from Healthy
#       |-> Module Set up
#       |-> Task 1. Possible Transitions from Healthy
#       |-> Task 2. Sample latent arrival times starting from Healthy
```

```
#           |-> Submodule  1.1: Transitions Healthy -> Sick
#           |-> Submodule  1.2: Transitions Healthy -> Dead
#       |-> Task 3: Predict the next state
#       |-> Task 4: Update important variables
#   |-> Module  2: Transitions from Sick
#       |-> Module Set up
#       |-> Task 1. Possible Transitions from Sick
#       |-> Task 2. Sample latent arrival times starting from Sick
#           |-> Submodule  2.1: Transitions Sick  ->  Healthy
#           |-> Submodule  2.2: Transitions Sick  ->  Sicker
#           |-> Submodule  2.3: Transitions Sick  ->  Dead
#       |-> Task 3: Predict the next state
#       |-> Task 4: Update important variables
#   |-> Module  3: Transitions from Sicker
#       |-> Module Set up
#       |-> Task 1. Possible Transitions from Sicker
#       |-> Task 2. Sample latent arrival times starting from Sicker
#           |-> Submodule  3.1: Transitions Sicker  ->  Dead
#       |-> Task 3: Predict the next state
#       |-> Task 4: Update important variables
#-----------------------------------------------------------------------------
--#
#  Module  1: Transitions from Healthy ----
#-----------------------------------------------------------------------------
--#
## Module Set up ----
# Objectives:
# 1. Generate  sub_tables to be used as inputs for each state-specific Module
## Inputs: dt_baseline, or dt_main;
## 1.1 At any given time determine how many people are in each non-absorbing
state using dt_main
## 1.2 Split the dt_main into k different sub-tables, one sub_dt for each of
k non-absorbing states
## 1.3 Store a list of state-specific sub-tables in a list, l_subsets_main_dt
.
#-----------------------------------------------------------------------------
--#

## Read dt
dt_main  <- copy(dt_baseline) # For first event

## Determine set of current states
v_current_state       <- unique(dt_main$from)
v_current_state_names <- paste("from", v_current_state, sep = "_")

## Subset dt_main based on origin state(s)
# For the purely progressive model and everyone starting at H,
# this step is trivial
l_subsets_main_dt <- list()
for( i in seq_along(v_current_state)){
```

```r
  l_subsets_main_dt[[i]] <-  as.data.table(dt_main[from == v_current_state[i]
, ])
}
names(l_subsets_main_dt) <- v_current_state_names

# ------------------------------------------- #
## Task 1. Possible Transitions from H ----
# Objective: Identify possible transitions out of the current state (H)
# Inputs: l_subsets_main_dt$from_H; m_Tr_r; dt_trans_keys,
# ------------------------------------------- #
# Temporary copy of dt_subset from H
dt_subset        <- l_subsets_main_dt$from_H
current_state    <- unique(dt_subset$from)     # current state
# Number of possible transitions out of Healthy
transition       <- as.numeric(m_Tr_r[current_state, m_Tr_r[current_state,] !
=0])
# Expand temp to have one row for each possible transition for each ID
ID               <- unique(dt_subset$ID)# unique IDs
setkey(dt_subset, ID)
## CJ is like expand.grid  for data.tables
dt_subset_long   <- merge(dt_subset,CJ(ID, transition), all.x = F)
dt_subset_long[, c("from","to") := NULL]

## Assign "from" and "to" state using dt_trans_keys
# Indexed join in data.table using "on" sets the Key automatically
dt_subset_long   <- dt_subset_long[dt_trans_keys, on = "transition", nomatch
= 0L]
dt_subset_long   <- dt_subset_long[order(ID, transition)]

# -------------------------------------------- #
## Task 2: Sample latent Event-times ----
# -------------------------------------------- #
### Sub-module 1-1: Transition #1  (H -> S1) ----
# -------------------------------------------- #

# Objective: Sample time of occurrence for transition H-> S1
# for every person currently in H.
# Inputs: dt_subset_long; r_HS1
# -------------------------------------------- #
# Constant annual rate of becoming Sick when Healthy
# Sample from an exponential distribution
if(r_HS1 != 0){   # Case when the transition is allowed
  dt_subset_long[transition == 1, T_stop := rexp(.N, rate = r_HS1) + T_n]
} else {        # Case when transition is not allowed, assign infinite time
  dt_subset_long[transition == 1, T_stop := Inf]
}
# ------------------------------------------- #
### Sub-module 1-2: Transition #2  (H -> D) ----
# -------------------------------------------- #
```

```
# Objective: Sample time of occurrence for transition H-> D
# for every person currently in H.
# Inputs: obtain_probs_des; dt_time2death_probs;dt_time2death_probs;
# dt_bckgrd_mortality; dt_subset_long
# ------------------------------------------- #
# Obtain vector of probabilities of dying from life table
dt_time2death_probs <- as.data.table(obtain_probs_des(dt_bckgrd_mortality))
# Only keep  rows for which the transition is possible,i.e., trans #2
dt_trans2           <- dt_subset_long[transition == 2]

# Set key in dt_trans2
setkey(dt_trans2, Age, Sex)
# Set key in dt_time2death_probs
setkey(dt_time2death_probs, Age, Sex)
# Indexed merge (all.x == T performs left-join)
dt_2sample_time2death <- merge(dt_trans2[, Age := ceiling(Age)], dt_time2deat
h_probs, all.x = TRUE)

# Generate a matrix of probabilities from rows that matched and merged.
m_probs <- as.matrix(dt_2sample_time2death[, .SD, .SDcols = patterns("^DP")])

# Sample time to death using nonparametric sampling, using `nps_nhppp` with a
uniform correction
dt_trans2[, T_stop := nps_nhppp(m_probs = m_probs, correction = "uniform")]

# Write into dt_subset_long
## guarantee that the order is correct when copying
setkey(dt_subset_long, ID, T_n)
setkey(dt_trans2, ID, T_n)
dt_subset_long[transition == 2, T_stop := dt_trans2$T_stop]

# --------------------------------------- #
## Task 3: Predict the Next Event from H ----
# Objectives: Determine which transition out of H occurs first
# Inputs: dt_subset_long
# --------------------------------------- #
# Create an indicator for minimum value in T_stop by ID
dt_subset_long[, status := as.numeric(T_stop == min(T_stop)), by = ID]
# Recover the inter-event time, tau
dt_subset_long[,tau := T_stop - T_start]
#---------------------------------------------------------------------------
--#
# Since everyone starts in the Healthy state and this is a progressive model
with
# no recurrence,we can skip the evaluation of the transitions out of
# the Sick and Sicker states in the first event.
#---------------------------------------------------------------------------
--#
```

```
# --------------------------------------- #
## Task 4: Update Important Variables ----
# Objectives: update important variables in the simulation
# including simulation time, age, event histories.
#  Updates differ  for the global event history, dt_event_history,
#  and for the data table used for the next event, dt_next_event
# Inputs: dt_subset_long, dt_event_history, dt_next_event
# --------------------------------------- #
# Update simulation time
dt_subset_long[status == T,T_n := T_stop]
# Update age
dt_subset_long[status == T,Age := round(T_n)]
# Update dt_event_history
dt_event_history  <- rbind(dt_event_history, dt_subset_long)
# Guarantee consistent ordering
setkey(dt_event_history, ID, T_n, Event_num, transition)
```

For a purely progressive model starting everyone in Healthy we skip Submodule 2 (transitions from Sick) and Submodule 3 (transitions from Sicker) in the first event, since everyone started in Healthy and after one iteration these same simulated people can either be in the Sick or Dead State. For determining the next event the only relevant people to keep track of in the simulation are those who transition to Sick.

```
#-----------------------------------------------------------------------------
--#
# Update dt_next_event
dt_next_event <- copy(dt_event_history)
setkey(dt_next_event,ID, T_n, Event_num, transition)
# Keep only transitions that occurred
dt_next_event <- dt_next_event[status == TRUE]
# only work with the last event, ignore previous events
dt_next_event <- dt_next_event[dt_next_event[, .I[Event_num == max(Event_num,
na.rm = TRUE)], by = ID]$V1]

# Handling Events beyond the time horizon
# Those who are alive beyond 100 years or older, assign death status
# (should me a negligible number with the baseline parameters)
dt_next_event[to != "D"  & Age >= time_horizon[2]-1, to :="D"]

# discard in the local data table those who died,
# for next event we only require those who remain alive
dt_next_event <- dt_next_event[to != "D"]

# Reset and update variables of interest
dt_next_event[,':=' (T_start = T_n,
                     tau = 0,
                     status  = NA,
                     Event_num = Event_num + 1,
                     from = to)]
dt_next_event[,':=' (to = NA,
```

```
                     T_stop = 0,
                     transition = NULL)]

# --------------------------------------------------------------------------------------------- #
# 2nd Event
# For a purely progressive model starting everyone in Healthy:
# For the next event after the first the only relevant people to
# keep track of in the simulation are those who transition to Sick
# --------------------------------------------------------------------------------------------- #
# Module 2: Transitions from Sick ----
# --------------------------------------------------------------------------------------------- #
## Module Set up ----
# Objectives of the Module:
# 1. Generate data subtables to be used as inputs for  each state-specific Module
## Inputs: dt_baseline, or dt_main;
## 1.1 At any given time determine how many people are in each non-absorbing state using dt_main
## 1.2 Split the dt_main into k different sub-tables, one sub_dt for each of k non-absorbing states
## 1.3 Store a list of state-specific sub-tables in a list, l_subsets_main_dt.
# --------------------------------------------------------------------------------------------- #
## Read dt_next_event
dt_main  <- dt_next_event
## Determine set of current states
v_current_state       <- unique(dt_main$from)
v_current_state_names <- paste("from", v_current_state, sep = "_")

## Subset dt_main based on origin state(s)
# For the purely progressive model and everyone starting at H,
# this step is trivial since everyone is in the Sick state or died after the first event.
l_subsets_main_dt <- list()
for( i in seq_along(v_current_state)){
  l_subsets_main_dt[[i]] <-  as.data.table(dt_main[from == v_current_state[i], ])
}
names(l_subsets_main_dt) <- v_current_state_names
# ----------------------------------- #
## Task 1: Possible Transitions from Sick ----
# Objective: Identify possible transitions out of the current state (Sick)
# Inputs: l_subsets_main_dt$from_S1; m_Tr_r; dt_trans_keys,
# ----------------------------------- #
# Temporary copy of dt_subset from S1
dt_subset        <- l_subsets_main_dt$from_S1
```

```
current_state    <- unique(dt_subset$from)     # current state
# Number of possible transitions out of S1
transition       <- as.numeric(m_Tr_r[current_state, m_Tr_r[current_state,] !
=0])
# Expand temp to have one row for each possible transition for each ID
ID               <- unique(dt_subset$ID)# unique IDs
setkey(dt_subset, ID)
## CJ is like expand.grid  for data.tables
dt_subset_long   <- merge(dt_subset,CJ(ID, transition), all.x = F)
dt_subset_long[, c("from","to") := NULL]

## Assign "from" and "to" state using dt_trans_keys
# Indexed join in data.table using "on" sets the Key automatically
dt_subset_long   <- dt_subset_long[dt_trans_keys, on = "transition", nomatch
= 0L]
dt_subset_long   <- dt_subset_long[order(ID, transition)]

# ----------------------------------- #
## Task 2: Sample latent Event-times ----
# ----------------------------------- #
# ----------------------------------- #
### Sub-module 2-1: Transition #3 (S1 -> H)  ----
# Objective: Sample time of occurrence for transition S1-> H
# for every person currently in S1
# Inputs: dt_subset_long; r_S1H
# ----------------------------------- #
# Enforce S1->H is not allowed (purely progressive)
# Sample from exponential distribution
r_S1H <- 0
if(r_S1H != 0){ # case when the transition is  allowed
  dt_subset_long[transition == 3, T_stop := rexp(.N, rate = r_S1H) + T_n]
}else {         # case when the transition is not  allowed
  dt_subset_long[transition == 3, T_stop := Inf]
}
# ----------------------------------- #
### Sub-module 2-2: Transition #4 (S1 -> S2) ----
# Objective: Sample time of occurrence for transition S1-> H
# for every person currently in S1
# Inputs: dt_subset_long; r_S1S2_scale_ph,r_S1S2_shape, hr_S1S2_trtB
# ----------------------------------- #
# State-residence time-dependent hazard of transition from Sick to Sicker
# Apply the treatment effect for strategy B or strategy AB
if (strategy == "B" || strategy == "AB"){
  r_S1S2_scale_ph <- r_S1S2_scale_ph * hr_S1S2_trtB
}
# Reparameterize to match accelerated failure time specification
r_S1S2_scale_aft <- r_S1S2_scale_ph ^(-1 / r_S1S2_shape)

# Sample inter-event time from Weibull distribution
if(r_S1S2_shape*r_S1S2_scale_aft != 0){  # case when the transition is  allow
```

```
ed
  dt_subset_long[transition == 4, T_stop := rweibull(.N, shape = r_S1S2_shape
, scale = r_S1S2_scale_aft) + T_n]
} else {                                # case when the transition is not  al
lowed
  dt_subset_long[transition == 4, T_stop := Inf]
}
# ----------------------------------- #
### Sub-module 2-2: Transition #5  (S1 -> D) ----
# Objective: Sample time of occurrence for transition S1-> D
# for every person currently in S1.
# Inputs: obtain_probs_des; dt_time2death_probs;dt_time2death_probs;
# dt_mortality_Sick; dt_subset_long
# ----------------------------------- #
# Obtain vector of probabilities of dying from life table
dt_time2death_probs_Sick              <- as.data.table(obtain_probs_des(dt_mo
rtality_Sick))

# Only keep  rows for which the transition is possible,i.e., trans #5
dt_trans5                             <- dt_subset_long[transition == 5]

# Set key in dt_trans5
setkey(dt_trans5, Age,Sex)
# Set key in dt_time2death_probs_Sick
setkey(dt_time2death_probs_Sick, Age,Sex)
# Indexed merge (all.x == T performs left-join)
dt_2sample_time2death_Sick          <- merge(dt_trans5[,Age := ceiling(Age)],
                                             dt_time2death_probs_Sick, all.x
= TRUE)

# Generate a matrix of probabilities from rows that matched and merged.
m_probs                             <- as.matrix(dt_2sample_time2death_Sick[,
.SD, .SDcols = patterns("^DP")])

# Sample time to death using nonparametric sampling, using `nps_nhppp` with a
uniform correction
dt_trans5[, T_stop := nps_nhppp(m_probs = m_probs, correction = "uniform")]
# Consistent ordering
setkey(dt_subset_long,ID,T_n)
setkey(dt_trans5,ID,T_n)
# Write into dt_subset_long
dt_subset_long[transition == 5, T_stop := dt_trans5$T_stop]
dt_subset_long[transition == 5]
# ----------------------------------- #
## Task 3: Predict the Next Event from Sick ----
# Objectives: Determine which transition out of S1 occurs first
# Inputs: dt_subset_long
# ----------------------------------- #
# Create an indicator for minimum value in T_stop by ID
```

```
dt_subset_long[, status := as.numeric(T_stop == min(T_stop)), by = ID]
# Recover the inter-event time, tau
dt_subset_long[,tau := T_stop - T_start]

# ----------------------------------- #
## Task 4: Update Important Variables ----
# Objectives: update important variables in the simulation
# including simulation time, age, event histories.
#  Updates differ  for the global event history, dt_event_history,
#  and for the data table used for the next event, dt_next_event
# Inputs: dt_subset_long, dt_event_history, dt_next_event
# ----------------------------------- #
# Update simulation time
dt_subset_long[status == T,T_n := T_stop]
# Update age
dt_subset_long[status == T,Age := round(T_n)]
# Update dt_event_history
dt_event_history  <- rbind(dt_event_history, dt_subset_long)
# Guarantee consistent ordering
setkey(dt_event_history, ID, T_n, Event_num, transition)

# Update dt_next_event
dt_next_event <- copy(dt_event_history)
setkey(dt_next_event,ID, T_n, Event_num, transition)
# Keep only transitions that occurred
dt_next_event <- dt_next_event[status == TRUE]
# only work with the last event, ignore previous events
dt_next_event <- dt_next_event[dt_next_event[, .I[Event_num == max(Event_num,
na.rm = TRUE)], by = ID]$V1]

# Handling Events beyond the time horizon
# Those who are alive beyond 100 years or older, assign death status
dt_next_event[to != "D"  & Age >= time_horizon[2]-1, to :="D"]
# discard those who died for next event
dt_next_event <- dt_next_event[to != "D"]

# #Reset and update variables of interest
dt_next_event[,':=' (T_start = T_n,
                     tau = 0,
                     status  = NA,
                     Event_num = Event_num + 1,
                     from = to)]
dt_next_event[,':=' (to = NA,
                     T_stop = 0,
                     transition = NULL)]

# ---------------------------------------------------------------------------
------------- #
# 3rd Event
```

```
#-----------------------------------------------------------------------------
--#
# For a purely progressive model starting everyone in Healthy:
# we can skip Submodule 1 (transitions from H)
# and Submodule 3 (transitions from Sicker) in the second event.
# In a purely progressive model with everyone starting in Healthy ,
# after the first event, nobody in the simulation stays Healthy.
# In a purely progressive model with everyone starting in Healthy ,
# after the second event, nobody in the simulation stays in Healthy nor Sick.
#-----------------------------------------------------------------------------
--#
# Module 3: Transitions from Sicker ----
#-----------------------------------------------------------------------------
--#
## Module Set up ----
# Objectives of the Module:
# 1. Generate data subtables to be used as inputs for  each state-specific Mo
dule
## Inputs: dt_baseline, or dt_main;
## 1.1 At any given time determine how many people are in each non-absorbing
state using dt_main
## 1.2 Split the dt_main into k different sub-tables, one sub_dt for each of
k non-absorbing states
## 1.3 Store a list of state-specific sub-tables in a list, l_subsets_main_dt
.
# ----------------------------------------------------------------------------
------------- #
## Read dt_next_event
dt_main  <- dt_next_event
## Determine set of current states
v_current_state       <- unique(dt_main$from)
v_current_state_names <- paste("from", v_current_state, sep = "_")

## Subset dt_main based on origin state(s)
# For the purely progressive model and everyone starting at H,
# this step is trivial since everyone is in the Sicker state
l_subsets_main_dt <- list()
for( i in seq_along(v_current_state)){
  l_subsets_main_dt[[i]] <-  as.data.table(dt_main[from == v_current_state[i]
, ])
}
names(l_subsets_main_dt) <- v_current_state_names
# ----------------------------------- #
## Task 1: Possible Transitions from Sicker ----
# ----------------------------------- #
# Temporary copy of dt_subset from S2
dt_subset        <- l_subsets_main_dt$from_S2
current_state    <- unique(dt_subset$from)     # current state
# Number of possible transitions out of S2
transition       <- as.numeric(m_Tr_r[current_state, m_Tr_r[current_state,] !
```

```
=0])
# Expand temp to have one row for each possible transition for each ID
ID                 <- unique(dt_subset$ID)# unique IDs
setkey(dt_subset, ID)
## CJ is like expand.grid  for data.tables
dt_subset_long   <- merge(dt_subset,CJ(ID, transition), all.x = F)
dt_subset_long[, c("from","to") := NULL]

## Assign "from" and "to" state using dt_trans_keys
# Indexed join in data.table using "on" sets the Key automatically
dt_subset_long   <- dt_subset_long[dt_trans_keys, on = "transition", nomatch
= 0L]
dt_subset_long   <- dt_subset_long[order(ID, transition)]
# ----------------------------------- #
## Task 2: Sample latent Event-times -----
# ----------------------------------- #
### Submodule 3-1: Transition #6  (S2 -> D) ----
# Objective: Sample time of occurrence for transition S2-> D
# for every person currently in S2.
# Inputs: obtain_probs_des; dt_time2death_probs;dt_time2death_probs;
# dt_mortality_Sicker; dt_subset_long
# ----------------------------------- #
# Obtain vector of probabilities of dying from life table
dt_time2death_probs_Sicker              <- as.data.table(obtain_probs_des(dt_
mortality_Sicker))

# Only keep  rows for which the transition is possible,i.e., trans #5
dt_trans6                             <- dt_subset_long[transition == 6]

# Set key in dt_trans5
setkey(dt_trans6, Age,Sex)
# Set key in dt_time2death_probs_Sick
setkey(dt_time2death_probs_Sicker, Age,Sex)
# Indexed merge (all.x == T performs left-join)
dt_2sample_time2death_Sicker          <- merge(dt_trans6[,Age := ceiling(Age)
],
                                                       dt_time2death_probs_Sicker, al
l.x = TRUE)

# Generate a matrix of probabilities from rows that matched and merged.
m_probs                             <- as.matrix(dt_2sample_time2death_Sicker
[, .SD, .SDcols = patterns("^DP")])

# Sample time to death using nonparametric sampling, using `nps_nhppp` with a
uniform correction
dt_trans6[, T_stop := nps_nhppp(m_probs = m_probs, correction = "uniform")]
# Consistent ordering
setkey(dt_subset_long,ID,T_n)
setkey(dt_trans6,ID,T_n)
```

```r
# Write into dt_subset_long
dt_subset_long[transition == 6, T_stop := dt_trans6$T_stop]
# ------------------------------------ #
## Task 3: Predict the Next Event from Sicker ----
# Objectives: Determine which transition out of S2 occurs first
# Inputs: dt_subset_long
# ------------------------------------ #
# Create an indicator for minimum value in T_stop by ID
dt_subset_long[, status := as.numeric(T_stop == min(T_stop)), by = ID]
# Recover the inter-event time, tau
dt_subset_long[,tau := T_stop - T_start]

# ------------------------------------ #
## Task 4: Update Important Variables ----
# Objectives: update important variables in the simulation
# including simulation time, age, event histories.
# Updates differ  for the global event history, dt_event_history,
# and for the data table used for the next event, dt_next_event
# Inputs: dt_subset_long, dt_event_history, dt_next_event
# ------------------------------------ #
# Update simulation time
dt_subset_long[status == T,T_n := T_stop]
# Update age
dt_subset_long[status == T,Age := round(T_n)]
# Update dt_event_history
dt_event_history  <- rbind(dt_event_history, dt_subset_long)
# Guarantee consistent ordering
setkey(dt_event_history, ID, T_n, Event_num, transition)

# ----#
# Quick Validation ----
# ---- #
# Verify everyone follows progression H->S1->S2->D, or dies in between
dt_event_history[, from_to := paste(from, to, sep = "_")]
table(dt_event_history[status==TRUE]$from_to)
# sum of all who die must equal n_sim
(table(dt_event_history[status==TRUE]$from_to)[1]+
  table(dt_event_history[status==TRUE]$from_to)[3]+
  table(dt_event_history[status==TRUE]$from_to)[5]) == n_sim
# Note some might have reached the time-horizon before dying,
# in this example it should be a negligible number
```

#### 2. DES Implementation of a Sick Sicker Model with Recursion to Healthy

```
#------------------------------------------------------------------------------
--#
#* To program this tutorial we used:
#* R version 4.2.1 (2022-06-23)
#* Platform: aarch64-apple-darwin20 (64-bit)
#* Running under: Mac OS 12.5
#* RStudio: Version 2022.07.1+554
#------------------------------------------------------------------------------
--#
# Description ----

#* This code implements a Discrete Event Simulation (DES) of a Sick-Sicker mo
del
#* with recurrence.
#* Structure:
#* Initial parameters stored in a list , `l_params`.
#* Modules are wrapped in the function `sim_next_event()`
#* which updates the `dt_next_event` and `dt_event_history`
#* The function `sim_next_event()` follows a modular structure :

# |-> Current Event Set up
#   |-> Module  1: Transitions from Healthy
#       |-> Module Set up
#       |-> Task 1. Possible Transitions from Healthy
#       |-> Task 2. Sample latent arrival times starting from Healthy
#           |-> Submodule  1.1: Transitions Healthy -> Sick
#           |-> Submodule  1.2: Transitions Healthy -> Dead
#       |-> Task 3: Predict the next state
#       |-> Task 4: Update important variables
#   |-> Module  2: Transitions from Sick
#       |-> Module Set up
#       |-> Task 1. Possible Transitions from Sick
#       |-> Task 2. Sample latent arrival times starting from Sick
#           |-> Submodule  2.1: Transitions Sick  ->  Healthy
#           |-> Submodule  2.2: Transitions Sick  ->  Sicker
#           |-> Submodule  2.3: Transitions Sick  ->  Dead
#       |-> Task 3: Predict the next state
#       |-> Task 4: Update important variables
#   |-> Module  3: Transitions from Sicker
#       |-> Module Set up
#       |-> Task 1. Possible Transitions from Sicker
#       |-> Task 2. Sample latent arrival times starting from Sicker
#           |-> Submodule  3.1: Transitions Sicker  ->  Dead
#       |-> Task 3: Predict the next state
#       |-> Task 4: Update important variables
# |-> Next Event Set up
```

```r
#* A while loop executes  `sim_next_event()`  until everyone in the simulatio
n has died
#* Complexity included in the model:
#* Time Dependence: the DES incorporate two types of type dependency
#* A. Simulation time-dependence (Age specific mortality)
#* B. State residency time-dependence (Transition S1 -> S2)
#*Treatment effect acting on transition rate S1->S2
#-----------------------------------------------------------------------------
--#
# Initial setup ----
rm(list = ls())    # remove any variables in R's memory
gc()               # clean working memory
#-----------------------------------------------------------------------------
--#
## Load packages ----
library("data.table"  )
library("dplyr"       )

## Load supplementary functions ----
source("R/DES_functions.R") # DES specific
source("R/Functions.R")     # General utility functions

#-----------------------------------------------------------------------------
--#
# Initialize Parameters ----
#* We want to pass parameters seamlessly through our function sim_next_event(
),
#* for this we can declare initial parameters in a list. Here we pass them th
rough
#* a function `init_param` that will return such list `l_params` and also inc
lude the baseline data table
#* `dt_baseline` as an object of that list, now named `dt_next_event`.

l_params <- init_params(
  # Sim. params
  # Time horizon
  time_init        = 25    ,
  time_end         = 100   ,
  # Sim. sample size
  n_sim            = 1e5   ,

  # dt_baseline information
  ## initial age
  age_init         = 25    ,
  ## sex group
  Sex_grp          = "Total",
  ## initial state
  state_init       = "H"   ,
```

```
  # State Space
  v_states          = c("H",                                                  # He
althy (H)
                        "S1",                                                 # Si
ck (S1)
                        "S2",                                                 # Si
cker (S2)
                        "D"),                                                 # De
ad (D)

  # Indexed Adjacency Matrix
  #* Zero entries indicate no transition between pairs of states,
  #* Nonzero positive entries index possible transitions in arbitrary order
  m_Tr_r            = matrix(c(0,1,0,2,
                               3,0,4,5,
                               0,0,0,6,
                               0,0,0,0),
                             nrow = 4, ncol = 4, byrow = TRUE,
                             dimnames = list(c("H", "S1","S2","D"),
                                             c("H", "S1","S2","D"))),
  # Transition Rates
  ## Mortality
  #* we specify subfolder in project to load Life Tables
  lifetable         = "Inputs/all_cause_mortality.Rda",
  ## Hazard Ratio while being S1
  hr_S1             =    3,
  ## Hazard Ratio while being S2
  hr_S2             =    10,

  ## Other transitions
  ## H  -> S1, constant rate ~Exp(r_HS1)
  r_HS1             =    0.15,
  ## S1 -> H,  constant rate ~Exp(r_S1H)
  r_S1H             =    0.5,
  ## S1 -> S2, state-residency time dependent ~ Weibull(r_S1S2_scale_ph, r_S1
S2_shape)
  ##* PH parameterization
  r_S1S2_scale_ph   =    0.08,
  r_S1S2_shape      =    1.1,

  # Strategy
  #* Can be any of "SoC", "A", "B", "AB".
  #* Only B and AB, affect transitions rates,
  strategy          =    "SoC",

  # Tx Effects that affect state-transitions
  #* Effectiveness of strategy B hazard ratio
  #* of becoming Sicker when Sick under treatment B
```

```
  hr_S1S2_trtB = 0.6
)

#-----------------------------------------------------------------------------
--#
# Simulation with Recurrence  ----
# ----------------------------------- #
# Initialize dt_next_event
dt_next_event      <- data.table()
# Initialize long-format dt_event_history
dt_event_history   <- data.table()
# Set seed for reproducibility
n_seed             <- 2
set.seed(n_seed)
# Max. number of events before triggering warning
# e.g., if more than 1,000 have occurred, trigger a warning message
max_num_events <- 1e3

#-----------------------------------------------------------------------------
--#
# Run while loop until everyone in the simulation has died
ptime             <- system.time({
  while(
    # stopping criteria: each row in dt_next_event represents
    # the number of people still alive in the simulation
    nrow(l_params$dt_next_event) >0
  ){

    # Function updates next event for everyone who is still alive
    l_params$dt_next_event <- sim_next_event(l_params)

    # Warning:
    if(unique(length(l_params$sim_next_event$Event_num)) > max_num_events){
      warning(
        paste0("Within a simulation time-horizon of"
               ,l_params$time_horizon[2]-l_params$time_horizon[1], "years,
                the max number of events (i.e., transitions between health
                states) for at least one person in the simulation has
                exceeded", max_num_events,".Reconsider if the simulation
                should continue since these dynamics might not reflect
                what you intended."
        ))
    }
  }
})[3]
ptime/60
ptime

#-----------------------------------------------------------------------------
```

```
--#
# Explore a single trajectory ----

# Output in long format
head(dt_event_history)
# Subset to events that occurred
head(dt_event_history[status == TRUE])

# Single trajectory
dt_event_history[status == TRUE & ID == 3]

# With the current setup
# Sim person with ID:3, experienced the following path:
# H->S1->H->S1->S2->D (read from cols "from" and "to")
# Events occurred at the following times:
# 25.97768 -> 26.88503 -> 28.52374 -> 29.20695 -> 76.18308 (read from col "T_
stop")
# Dwell times at each alive states:
# H(0.9776845), S1(0.9073458), H(1.6387102), S1(0.6832138), S2(46.9761233)  (
read from col "tau")
#------------------------------------------------------------------------------
--#
```

### 3. Module A: Cost-effectiveness Analysis of a Sick Sicker Model with Recursion

This Module executes a Discrete Event Simulation (DES) of a Sick-Sicker model under four different strategies and computes a cost-effectiveness analysis.

The four strategies are:

- Standard of Care (SoC): best available care for the patients with the disease. This scenario reflects the natural history of the disease progression.
- Strategy A: treatment A is given to patients in the Sick and Sicker states, but only improves the quality of life of those in the Sick state.
- Strategy B: treatment B is given to all sick patients and reduces disease progression from the Sick to Sicker state.
- Strategy AB: This strategy combines treatment A and treatment B. The disease progression is reduced, and individuals in the Sick state have an improved quality of life.

CEA Outcomes:

- ICERs

```
#-----------------------------------------------------------------------------
--#
# Initial setup ----
rm(list = ls())    # remove any variables in R's memory
gc()               # clean working memory
#-----------------------------------------------------------------------------
--#
## Load packages ----
library("data.table"  )
library("dplyr"       )

## Load supplementary functions ----
source("R/DES_functions.R") # DES specific
source("R/Functions.R")     # General utility functions
#-----------------------------------------------------------------------------
--#
# Initialize Parameters ----
#* We want to pass parameters seamlessly through our function sim_next_event(
),
#* for this we can declare initial parameters in a list. Here we pass them th
rough
#* a function `init_param` that will return such list `l_params` and also inc
lude the baseline data table
#* `dt_baseline` as an object of that list, now named `dt_next_event`.
```

```
l_params <- init_params(
  # Sim. params
  # Time horizon
  time_init         = 25    ,
  time_end          = 100   ,
  # Sim. sample size
  n_sim             = 1e5   ,

  # dt_baseline information
  ## initial age
  age_init          = 25    ,
  ## sex group
  Sex_grp           = "Total",
  ## initial state
  state_init        = "H"   ,

  # State Space
  v_states          = c("H",                                                  # He
althy (H)
                        "S1",                                                 # Si
ck (S1)
                        "S2",                                                 # Si
cker (S2)
                        "D"),                                                 # De
ad (D)

  # Indexed Adjacency Matrix
  #* Zero entries indicate no transition between pairs of states,
  #* Nonzero positive entries index possible transitions in arbitrary order
  m_Tr_r            = matrix(c(0,1,0,2,
                               3,0,4,5,
                               0,0,0,6,
                               0,0,0,0),
                             nrow = 4, ncol = 4, byrow = TRUE,
                             dimnames = list(c("H", "S1","S2","D"),
                                             c("H", "S1","S2","D"))),
  # Transition Rates
  ## Mortality
  #* we specify subfolder in project to load Life Tables
  lifetable         = "Inputs/all_cause_mortality.Rda",
  ## Hazard Ratio while being S1
  hr_S1             =    3,
  ## Hazard Ratio while being S2
  hr_S2             =    10,

  ## Other transitions
  ## H  -> S1, constant rate ~Exp(r_HS1)
  r_HS1             =    0.15,
  ## S1 -> H,  constant rate ~Exp(r_S1H)
```

```r
  r_S1H             =    0.5,
  ## S1 -> S2, state-residency time dependent ~ Weibull(r_S1S2_scale_ph, r_S1
S2_shape)
  ##* PH parameterization
  r_S1S2_scale_ph  =    0.08,
  r_S1S2_shape     =    1.1,

  # Strategy
  #* Can be any of "SoC", "A", "B", "AB".
  #* Only B and AB, affect transitions rates,
  strategy          =    "SoC",

  # Tx Effects that affect state-transitions
  #* Effectiveness of strategy B hazard ratio
  #* of becoming Sicker when Sick under treatment B
  hr_S1S2_trtB = 0.6
)
str(l_params)
#  Reference l_params
l_params_ref          <- l_params
#------------------------------------------------------------------------------
--#
## Run Simulation for all Strategies
#------------------------------------------------------------------------------
--#
v_strategies_names <- c( "SoC", "A", "B", "AB" )
#* since the effect on transition dynamics from SoC == A, and B == AB,
#* we can just execute the simulation for "SOC" and "B"
v_strategies       <- c("SoC","B")
# Set seed for reproducibility
n_seed             <- 2
#Initialize list of event histories
l_event_history_strategies   <- list()

# For-loop running simulation under each strategy
for(i in seq_along(v_strategies)){
  # Initialize dt_event_history for each strategy
  dt_event_history    <- data.table()
  # Reset initial params for each iteration
  l_params            <- l_params_ref
  # Update strategy
  l_params$strategy   <- v_strategies[i]
  # Common random numbers
  set.seed(n_seed)
  # Run simulation under i^th strategy
  while(nrow(l_params$dt_next_event) > 0){
    l_params$dt_next_event <- sim_next_event(l_params)
  }
  # Store dt_event_history for i^th strategy
  l_event_history_strategies[[i]] <-as.data.table(dt_event_history)
```

```r
}
names(l_event_history_strategies) <- v_strategies

# Duplicating event histories for equivalent strategies
# "SoC" and "A"
l_event_history_strategies$A  <- l_event_history_strategies$SoC
# "B" and "AB"
l_event_history_strategies$AB <- l_event_history_strategies $B
#View(l_event_history_strategies)

#-----------------------------------------------------------------------------
--#
# Cost-Effectiveness Analysis (CEA) ----
#-----------------------------------------------------------------------------
--#
# CEA Parameters ----
# -------------------------- #

#* We store the parameters necessary for the cost-effectiveness analysis
#* on a list `l_cea_params`.
#* Note we store the list of event histories in `l_cea_params`.

l_cea_params<- list(
  # list of event histories for each strategy
  l_event_history_strategies = l_event_history_strategies,

  ## Discounting factors ----
  d_c       = 0.03, # annual discount rate for costs
  d_e       = 0.03, # annual discount rate for QALYs

  ## State rewards ----
  ### Costs ----
  c_H       = 2000  , # annual cost of being Healthy
  c_S1      = 4000  , # annual cost of being Sick
  c_S2      = 15000 , # annual cost of being Sicker
  c_D       = 0     , # annual cost of being dead
  dc_trtA   = 12000 , # annual cost of receiving treatment A
  dc_trtB   = 13000 , # annual cost of receiving treatment B
  ### Utilities ----
  u_H       = 1     , # annual utility of being Healthy
  u_S1      = 0.75  , # annual utility of being Sick
  u_S2      = 0.5   , # annual utility of being Sicker
  u_D       = 0     , # annual utility of being dead
  u_trtA    = 0.95  , # annual utility when receiving treatment A
  du_trtA   = 0.95-0.75,
  ### Transition rewards ----
  du_HS1    = -0.01 , # disutility when transitioning from Healthy to Sick
  dc_HS1    = 1000  , # increase in cost when transitioning from Healthy to Si
```

```
ck
  dc_D      = 2000    # increase in cost when dying

)

# ---------------------------------- #
# Compute CEA ----
# ---------------------------------- #
## ICERs ----
#* `cea_fn` computes the cea analysis given a list of event histories,
#* that correspond to a strategy. The details of the function are explained i
n the
#* manuscript subsection: Post-processing Module A: Compute a Cost-effectiven
ess Analysis
#* The output is the result of a competing choice analysis

table_cea<- cea_fn(l_cea_params = l_cea_params)
table_cea
```

#### 4. Module B: Epidemiological Outcomes of a Sick Sicker Model with Recursion

```
#------------------------------------------------------------------------------
--#
#* To program this tutorial we used:
#* R version 4.2.1 (2022-06-23)
#* Platform: aarch64-apple-darwin20 (64-bit)
#* Running under: Mac OS 12.5
#* RStudio: Version 2022.07.1+554
#------------------------------------------------------------------------------
--#
# Description ----
#* This Module executes a Discrete Event Simulation (DES) of a Sick-Sicker mo
del
#* under four different strategies and computes a relevant epidemiological ou
tcomes
#* for the simulated cohort.
#* Strategies are:
#*  - Standard of Care (SoC): best available care for the patients with the
#*    disease. This scenario reflects the natural history of the disease
#*    progression.
#*  - Strategy A: treatment A is given to patients in the Sick and Sicker sta
tes,
#*    but only improves the quality of life of those in the Sick state.
#*  - Strategy B: treatment B is given to all sick patients and reduces disea
se
#*    progression from the Sick to Sicker state.
#*  - Strategy AB: This strategy combines treatment A and treatment B. The di
sease
#*    progression is reduced, and individuals in the Sick state have an impro
ved
#*    quality of life.
#* Epidemiological Outcomes:
#*  - Survival: Probability of surviving up to time t
#*  - Prevalence of Disease: total number of simulated people
#*    with disease (i.e., in Sick or Sicker states) divided by the total numb
er  of
#*    alive population at the beginning of the simulation
#*  - Average Dwell Times: time spent in a given alive state  on average over
the
#*    follow-up period.
#*  - Distribution of Average Dwell Times: across the simulated cohort, the d
istribution
#*    of time the people spent on a given alive state over the follow-up peri
od on average.
#*
#------------------------------------------------------------------------------
--#
# Initial setup ----
```

```
rm(list = ls())    # remove any variables in R's memory
gc()               # clean working memory
#----------------------------------------------------------------------------
--#
## Load packages ----
library("data.table"  )
library("dplyr"       )

## Load supplementary functions ----
source("R/DES_functions.R") # DES specific
source("R/Functions.R")     # General utility functions
#----------------------------------------------------------------------------
--#
# Initialize Parameters ----
#* We want to pass parameters seamlessly through our function sim_next_event(
),
#* for this we can declare initial parameters in a list. Here we pass them th
rough
#* a function `init_param` that will return such list `l_params` and also inc
lude the baseline data table
#* `dt_baseline` as an object of that list, now named `dt_next_event`.

l_params <- init_params(
  # Sim. params
  # Time horizon
  time_init        = 25    ,
  time_end         = 100   ,
  # Sim. sample size
  n_sim            = 1e3   ,

  # dt_baseline information
  ## initial age
  age_init         = 25    ,
  ## sex group
  Sex_grp          = "Total",
  ## initial state
  state_init       = "H"   ,

  # State Space
  v_states         = c("H",                                           # He
althy (H)
                       "S1",                                          # Si
ck (S1)
                       "S2",                                          # Si
cker (S2)
                       "D"),                                          # De
ad (D)

  # Indexed Adjacency Matrix
```

```
  #* Zero entries indicate no transition between pairs of states,
  #* Nonzero positive entries index possible transitions in arbitrary order
  m_Tr_r           = matrix(c(0,1,0,2,
                              3,0,4,5,
                              0,0,0,6,
                              0,0,0,0),
                            nrow = 4, ncol = 4, byrow = TRUE,
                            dimnames = list(c("H", "S1","S2","D"),
                                            c("H", "S1","S2","D"))),
  # Transition Rates
  ## Mortality
  #* we specify subfolder in project to load Life Tables
  lifetable        = "Inputs/all_cause_mortality.Rda",
  ## Hazard Ratio while being S1
  hr_S1            =    3,
  ## Hazard Ratio while being S2
  hr_S2            =    10,

  ## Other transitions
  ## H  -> S1, constant rate ~Exp(r_HS1)
  r_HS1            =    0.15,
  ## S1 -> H,  constant rate ~Exp(r_S1H)
  r_S1H            =    0.5,
  ## S1 -> S2, state-residency time dependent ~ Weibull(r_S1S2_scale_ph, r_S1
S2_shape)
  ##* PH parameterization
  r_S1S2_scale_ph  =    0.08,
  r_S1S2_shape     =    1.1,

  # Strategy
  #* Can be any of "SoC", "A", "B", "AB".
  #* Only B and AB, affect transitions rates,
  strategy         =    "SoC",

  # Tx Effects that affect state-transitions
  #* Effectiveness of strategy B hazard ratio
  #* of becoming Sicker when Sick under treatment B
  hr_S1S2_trtB = 0.6
)
str(l_params)
#  Reference l_params
l_params_ref          <- l_params
#-----------------------------------------------------------------------------
--#
## Run Simulation for all Strategies
#-----------------------------------------------------------------------------
--#
v_strategies_names <- c( "SoC", "A", "B", "AB" )
#* since the effect on transition dynamics from SoC == A, and B == AB,
#* we can just execute the simulation for "SOC" and "B"
```

```
v_strategies       <- c("SoC","B")
# Set seed for reproducibility
n_seed             <- 2
#Initialize list of event histories
l_event_history_strategies   <- list()

# For-loop running simulation under each strategy
for(i in seq_along(v_strategies)){
  # Initialize dt_event_history for each strategy
  dt_event_history    <- data.table()
  # Reset initial params for each iteration
  l_params            <- l_params_ref
  # Update strategy
  l_params$strategy   <- v_strategies[i]
  # Common random numbers
  set.seed(n_seed)
  # Run simulation under i^th strategy
  while(nrow(l_params$dt_next_event) > 0){
    l_params$dt_next_event <- sim_next_event(l_params)
  }
  # Store dt_event_history for i^th strategy
  l_event_history_strategies[[i]] <-as.data.table(dt_event_history)
}
names(l_event_history_strategies) <- v_strategies

# Duplicating event histories for equivalent strategies
# "SoC" and "A"
l_event_history_strategies$A  <- l_event_history_strategies$SoC
# "B" and "AB"
l_event_history_strategies$AB <- l_event_history_strategies $B
#View(l_event_history_strategies)

#------------------------------------------------------------------------------
--#
#  Epidemiological Outcomes ----
#------------------------------------------------------------------------------
--#
## Compute Epi Outcomes ----
#* Function `epi_fn` takes as input the list of the event histories under eac
h
#* strategy and returns the epidemiological outcomes on a list.
#* a. '$dt_dwell' and '$dt_dwell_pop' contain dwell times by health state, su
mmarized at the individual level or by strategy
#* b. '$l_dt_trace_strategies' contains for each strategy
#*    - state occupancy over time,
#*    - survival probabilities over time, and
#*    - disease prevalence over time

l_epi_outcomes <- epi_fn(l_event_history_strategies = l_event_history_strateg
```

```
ies)
View(l_epi_outcomes)
```

#### 5. Module C: Probabilistic Sensitivity Analysis of a Sick Sicker Model with Recursion

```
#------------------------------------------------------------------------------
--#
#------------------------------------------------------------------------------
--#
#* To program this tutorial we used:
#* R version 4.2.1 (2022-06-23)
#* Platform: aarch64-apple-darwin20 (64-bit)
#* Running under: Mac OS 12.5
#* RStudio: Version 2022.07.1+554
#------------------------------------------------------------------------------
--#
# Description ----

#* This code implements a Probabilistic  Analysis of a
#* CEA of the Sick-Sicker model under four strategies
#* The simulation is implemented as a Discrete Event Simulation (DES)
#* The four strategies are the following
#* - Standard of Care (SoC): best available care for the patients with the
#*   disease. This scenario reflects the natural history of the disease
#*   progression.
#* - Strategy A: treatment A is given to patients in the Sick and Sicker stat
es,
#*   but only improves the quality of life of those in the Sick state.
#* - Strategy B: treatment B is given to all sick patients and reduces diseas
e
#*   progression from the Sick to Sicker state.
#* - Strategy AB: This strategy combines treatment A and treatment B. The dis
ease
#*   progression is reduced, and individuals in the Sick state have an improv
ed
#*   quality of life.
#* Probabilistic Analysis-based Economic Outcomes
#* - Cost-effectiveness scatter plot
#* - Cost-effectiveness acceptability curves (CEACs) and frontier (CEAF)
#* - Expected Loss Curves (ELCs)
#* - Expected value of perfect information (EVPI)
#------------------------------------------------------------------------------
--#
# Initial setup ----
rm(list = ls())    # remove any variables in R's memory
gc()               # clean working memory
#------------------------------------------------------------------------------
--#
## Load packages ----
library("data.table"  )
library("dplyr"       )
library("doParallel"  )
```

```
library("parallel"    )
library("foreach"     )
library("MethylCapSig")

## Load supplementary functions ----
source("R/DES_functions.R") # DES specific
source("R/Functions.R")     # General utility functions
# PSA functions from previous tutorials
# (with permission from Dr. Alarid-Escudero)
# source("R/Functions_cSTM_time_dep_state_residence.R")
#-----------------------------------------------------------------------------
--#

# Initialize Sim. Model Parameters ----
#* We want to pass parameters seamlessly through our function sim_next_event(
),
#* for this we can declare initial parameters in a list. Here we pass them th
rough
#* a function `init_param` that will return such list `l_params` and also inc
lude
#* the baseline data table `dt_baseline` as an object of that list, now named
`dt_next_event`.

l_params <- init_params(
  # Sim. params
  # Time horizon
  time_init         = 25    ,
  time_end          = 100   ,
  # Sim. sample size
  n_sim             = 1e5   ,

  # dt_baseline information
  ## initial age
  age_init          = 25    ,
  ## sex group
  Sex_grp           = "Total",
  ## initial state
  state_init        = "H"   ,

  # State Space
  v_states          = c("H",                                   # Healthy (H)
                        "S1",                                  # Sick (S1)
                        "S2",                                  # Sicker (S2)
                        "D"),                                  # Dead (D)

  # Indexed Adjacency Matrix
  #* Zero entries indicate no transition between pairs of states,
  #* Nonzero positive entries index possible transitions in arbitrary order
  m_Tr_r            = matrix(c(0,1,0,2,
```

```
                              3,0,4,5,
                              0,0,0,6,
                              0,0,0,0),
                            nrow = 4, ncol = 4, byrow = TRUE,
                            dimnames = list(c("H", "S1","S2","D"),
                                            c("H", "S1","S2","D"))),
  # Transition Rates
  ## Mortality
  #* we specify subfolder in project to load Life Tables
  lifetable         = "Inputs/all_cause_mortality.Rda",
  ## Hazard Ratio while being S1
  hr_S1             =    3,
  ## Hazard Ratio while being S2
  hr_S2             =    10,

  ## Other transitions
  ## H  -> S1, constant rate ~Exp(r_HS1)
  r_HS1             =    0.15,
  ## S1 -> H,  constant rate ~Exp(r_S1H)
  r_S1H             =    0.5,
  ## S1 -> S2, state-residency time dependent ~ Weibull(r_S1S2_scale_ph, r_S1
S2_shape)
  ##* PH parameterization
  r_S1S2_scale_ph   =    0.08,
  r_S1S2_shape      =    1.1,

  # Strategy
  #* Can be any of "SoC", "A", "B", "AB".
  #* Only B and AB, affect transitions rates,
  strategy          =    "SoC",

  # Tx Effects that affect state-transitions
  #* Effectiveness of strategy B hazard ratio
  #* of becoming Sicker when Sick under treatment B
  hr_S1S2_trtB = 0.6
)
str(l_params)
#  Reference l_params
l_params_ref          <- l_params
# Store the parameter names into a vector
v_names_params <- names(l_params_ref)

# ------------------------------------------------------------------------ #
# Initialize Economic Parameters ----
l_cea_params<- list(
  # # list of event histories for each strategy
  l_event_history_strategies = NA,

  ## Discounting factors ----
```

```
  d_c      = 0.03, # annual discount rate for costs
  d_e      = 0.03, # annual discount rate for QALYs

  ## State rewards ----
  ### Costs ----
  c_H      = 2000  , # annual cost of being Healthy
  c_S1     = 4000  , # annual cost of being Sick
  c_S2     = 15000 , # annual cost of being Sicker
  c_D      = 0     , # annual cost of being dead
  dc_trtA  = 12000 , # annual cost of receiving treatment A
  dc_trtB  = 13000 , # annual cost of receiving treatment B
  ### Utilities ----
  u_H      = 1     , # annual utility of being Healthy
  u_S1     = 0.75  , # annual utility of being Sick
  u_S2     = 0.5   , # annual utility of being Sicker
  u_D      = 0     , # annual utility of being dead
  u_trtA   = 0.95  , # annual utility when receiving treatment A
  du_trtA  = 0.95-0.75,
  ### Transition rewards ----
  du_HS1   = -0.01 , # disutility when transitioning from Healthy to Sick
  dc_HS1   = 1000  , # increase in cost when transitioning from Healthy to Si
ck
  dc_D     = 2000    # increase in cost when dying
)

#  Reference l_params
l_cea_params_ref          <- l_cea_params

# Store the parameter names into a vector
v_names_cea_params <- names(l_cea_params_ref)

#-----------------------------------------------------------------------------
--#
#  Probabilistic Analysis input Dataset ----
n_sim_psa   <- 1e3

v_names_str <- c("Standard of care",      # store the strategy names
                 "Strategy A",
                 "Strategy B",
                 "Strategy AB")
n_str       <- length(v_names_str)        # number of strategies

# Generate PSA Input
dt_psa_input <- as.data.table(generate_psa_params_DES(n_sim = n_sim_psa))
# First six observations
# head(dt_psa_input)
#-----------------------------------------------------------------------------
--#
#------------------ #
```

```
# Run Probabilistic Analysis  -----
#------------------- #
## Initialize data.frames with Probabilistic Analysis  output  ----
### data.frame of costs
df_c <- as.data.frame(
  matrix(0,
         nrow = n_sim_psa,
         ncol = n_str))
colnames(df_c) <- v_names_str

### data.frame of effectiveness
df_e <- as.data.frame(
  matrix(0,
         nrow = n_sim_psa,
         ncol = n_str))
colnames(df_e) <- v_names_str

###  data.frame of NMB
df_nmb <- as.data.frame(
  matrix(0,
         nrow = n_sim_psa,
         ncol = n_str))
colnames(df_nmb) <- v_names_str

v_strategies_names <- c( "SoC", "A", "B", "AB" )
#* since the effect on transition dynamics from SoC == A, and B == AB,
#* we can just execute the simulation for "SOC" and "B"
v_strategies       <- c("SoC","B")
# Set seed for reproducibility
n_seed             <- 2
# ------------------------------------------------------------------------ #
# ------------------------------------------------------------------------ #

# WARNING: DO NOT RUN THE NEXT CHUNK UNLESS YOU HAVE ENOUGH COMPUTING RESOURC
ES

# ------------------------------------------------------------------------ #
# ------------------------------------------------------------------------ #
strategies <- v_strategies
## Non-distributed Implementation ----
# for (i in 1:n_sim_psa) {
#   print(paste("Init_PSA_iteration",i, sep = "_"))
#   l_params                  <- update_param_list(l_params_ref, dt_psa_input
[i,])
#   l_cea_params              <- update_param_list(l_cea_params_ref, dt_psa_i
nput[i,])
#   # A. Full Histories
#   l_event_history_strategies        <- list()
#   # Loop over strategies
```

```
#   for(j in seq_along(strategies)){
#     print(paste("Strategy",strategies[j],"iteration",i, sep = "_"))
#     l_params$strategy           <- strategies[j]
#     dt_event_history            <- data.table()
#     l_params$dt_next_event      <- l_params_ref$dt_next_event
#     # common random numbers for strategies
#     set.seed(n_seed)
#     # run simulation under i^th strategy
#     while(nrow(l_params$dt_next_event) >0){
#       l_params$dt_next_event          <- sim_next_event(l_params)
#     }
#     l_event_history_strategies[[j]]   <- as.data.table(dt_event_history)
#   }
#   # Store event-history for all strategies
#   l_event_history_strategies$A        <- l_event_history_strategies[[1]]
#   l_event_history_strategies$AB       <- l_event_history_strategies[[2]]
#   names(l_event_history_strategies)   <- c(strategies, "A","AB")
#
#   # Economics Outcomes
#   l_cea_params$l_event_history_strategies <- l_event_history_strategies
#   l_cea_PSA_iter                  <- cea_psa_fn(l_cea_params = l_cea_params
)
#   # PSA based outcomes
#   df_c[i, ]                       <- l_cea_PSA_iter$Cost
#   df_e[i, ]                       <- l_cea_PSA_iter$Effect
#   df_nmb[i,]                      <- l_cea_PSA_iter$NMB
#   print(paste("Fin_PSA_iteration",i, sep = "_"))
#   # # Display simulation progress
#   if (i/(n_sim_psa/100) == round(i/(n_sim_psa/100), 0)) { # display progres
s every 5%
#     cat('\r', paste(i/n_sim_psa * 100, "% done", sep = " "))
#   }
# }

# # ------------------------------------------------------------------------
#
# # ------------------------------------------------------------------------
#
# ## Distributed Implementation  ----
# Num. of Cores
 no_cores_all  <- parallel::detectCores()-1      # recommended when not shari
ng computing resources
# cores_90pct   <- as.integer(.90 * no_cores_all) # aggressive  when shared r
esources
 cores_85pct   <- as.integer(.85 * no_cores_all) # aggressive  when shared re
sources
# cores_75pct   <- as.integer(.75 * no_cores_all) # aggressive  when shared r
esources
# cores_50pct   <- as.integer(.5  * no_cores_all) # recommended when shared r
esources
```

```
# cores_25pct   <- as.integer(.25 * no_cores_all) # recommended when shared r
esources

#### Cluster  ----
os_info  <- Sys.info()
os       <- os_info[1]
no_cores <- cores_85pct

if (os == "Darwin") { # Apple Kernel
  # Initialize cluster object
  cl <- parallel::makeForkCluster(no_cores)
  # Register clusters
  doParallel::registerDoParallel(cl)
}
if (os == "Linux") {
  # Register clusters
  doMC::registerDoMC(cores = no_cores)
}
if (os == "Windows") {
  # Initialize cluster object
  cl <- parallel::makeCluster(no_cores)
  # Register clusters
  doParallel::registerDoParallel(cl)
}

#### Run distributed Probabilistic Analysis  ----
df_ce <- foreach::foreach(i = 1:n_sim_psa, .combine = rbind,
                          .packages = c("data.table","dplyr")
) %dopar% {
  #param lists
  l_params                <- update_param_list(l_params_ref, dt_psa_input[i
,])
  l_cea_params            <- update_param_list(l_cea_params_ref, dt_psa_inp
ut[i,])
  # A. Simulate Full Event Histories
  l_event_history_strategies              <- list()
  ## Loop over strategies
  for(j in seq_along(strategies)){
    #print(paste("Strategy",strategies[j],"iteration",j, sep = "_"))
    l_params$strategy             <- strategies[j]
    dt_event_history              <- data.table()
    l_params$dt_next_event        <- l_params_ref$dt_next_event
    ### common random numbers for strategies
    set.seed(n_seed)
    ### run simulation under i^th strategy
    while(nrow(l_params$dt_next_event) >0){
      l_params$dt_next_event              <- sim_next_event(l_params)
    }
```

```
    l_event_history_strategies[[j]]       <- as.data.table(dt_event_history)
  }
  ## Store event-history for all strategies
  l_event_history_strategies$A            <- l_event_history_strategies[[1]]
  l_event_history_strategies$AB           <- l_event_history_strategies[[2]]
  names(l_event_history_strategies)       <- c(strategies, "A","AB")

  # B. Compute Economic Outcomes
  l_cea_params$l_event_history_strategies <- l_event_history_strategies
  l_cea_PSA_iter                          <- cea_psa_fn(l_cea_params = l_cea_
params)
  df_ce                                   <- c(l_cea_PSA_iter$Cost, l_cea_PSA
_iter$Effect)
}
# Extract costs and effects from the PSA dataset
df_c <- df_ce[, 1:n_str]                # columns with costs
df_e <- df_ce[, (n_str + 1):(2*n_str)]  # columns with effects
# Stop clusters
parallel::stopCluster(cl)
# ------------------------------------------------------------------------ #
# Probabilistic Analysis  Data Object ----
# #* Function included in "R/Functions.R" The latest version can be found in
`dampack` package
l_psa <- make_psa_obj(cost          = df_c,
                      effectiveness = df_e,
                      parameters    = dt_psa_input,
                      strategies    = v_names_str)
l_psa$strategies            <- v_names_str
colnames(l_psa$effectiveness) <- v_names_str
colnames(l_psa$cost)          <- v_names_str
#View(l_psa)
# ---------------------------------- #
# ---------------------------------- #
```

#### 6. Auxiliary Module: Continuous Time Trace Matrix

```
#----------------------------------------------------------------------------
--#
#----------------------------------------------------------------------------
--#
#* To program this tutorial we used:
#* R version 4.2.1 (2022-06-23)
#* Platform: aarch64-apple-darwin20 (64-bit)
#* Running under: Mac OS 12.5
#* RStudio: Version 2022.07.1+554
#----------------------------------------------------------------------------
--#
# Description ----

#* In this Module we illustrate how to use functions
#* `cont_time_trace_sick_sicker`,
#* and  `discr_time_trace_sick_sicker`,
#* to transform the  output of the Discrete Event Simulation (DES) of
#* the Sick-Sicker model with or without recurrence into a continuous time
#* trace matrix or a discrete time approximation,
#* and how to produce a trace plot from the trace matrices.

#----------------------------------------------------------------------------
--#
# Initial setup ----
rm(list = ls())    # remove any variables in R's memory
gc()               # clean working memory
#----------------------------------------------------------------------------
--#
## Load packages ----
library("data.table"  )
library("dplyr"       )

## Load supplementary functions ----
source("R/DES_functions.R") # DES specific
source("R/Functions.R")     # General utility functions

#----------------------------------------------------------------------------
--#
# Initialize Parameters ----
#* We want to pass parameters seamlessly through our function sim_next_event(
),
#* for this we can declare initial parameters in a list. Here we pass them th
rough
#* a function `init_param` that will return such list `l_params` and also inc
lude the baseline data table
#* `dt_baseline` as an object of that list, now named `dt_next_event`.

l_params <- init_params(
```

```
  # Sim. params
  # Time horizon
  time_init         = 25    ,
  time_end          = 100   ,
  # Sim. sample size
  n_sim             = 1e3   ,

  # dt_baseline information
  ## initial age
  age_init          = 25    ,
  ## sex group
  Sex_grp           = "Total",
  ## initial state
  state_init        = "H"   ,

  # State Space
  v_states          = c("H",                                             # He
althy (H)
                        "S1",                                            # Si
ck (S1)
                        "S2",                                            # Si
cker (S2)
                        "D"),                                            # De
ad (D)

  # Indexed Adjacency Matrix
  #* Zero entries indicate no transition between pairs of states,
  #* Nonzero positive entries index possible transitions in arbitrary order
  m_Tr_r            = matrix(c(0,1,0,2,
                               3,0,4,5,
                               0,0,0,6,
                               0,0,0,0),
                             nrow = 4, ncol = 4, byrow = TRUE,
                             dimnames = list(c("H", "S1","S2","D"),
                                             c("H", "S1","S2","D"))),
  # Transition Rates
  ## Mortality
  #* we specify subfolder in project to load Life Tables
  lifetable         = "Inputs/all_cause_mortality.Rda",
  ## Hazard Ratio while being S1
  hr_S1             =    3,
  ## Hazard Ratio while being S2
  hr_S2             =    10,

  ## Other transitions
  ## H  -> S1, constant rate ~Exp(r_HS1)
  r_HS1             =    0.15,
  ## S1 -> H,  constant rate ~Exp(r_S1H)
  r_S1H             =    0.5,
```

```
  ## S1 -> S2, state-residency time dependent ~ Weibull(r_S1S2_scale_ph, r_S1
S2_shape)
  ##* PH parameterization
  r_S1S2_scale_ph  =    0.08,
  r_S1S2_shape     =    1.1,

  # Strategy
  #* Can be any of "SoC", "A", "B", "AB".
  #* Only B and AB, affect transitions rates,
  strategy         =    "SoC",

  # Tx Effects that affect state-transitions
  #* Effectiveness of strategy B hazard ratio
  #* of becoming Sicker when Sick under treatment B
  hr_S1S2_trtB = 0.6
)

#-----------------------------------------------------------------------------
--#
# Simulation with Recurrence  ----
# ----------------------------------- #
# Initialize dt_next_event
dt_next_event      <- data.table()
# Initialize long-format dt_event_history
dt_event_history   <- data.table()
# Set seed for reproducibility
n_seed             <- 2
set.seed(n_seed)
# Max. number of events before triggering warning
# e.g., if more than 1,000 have occurred, trigger a warning message
max_num_events <- 1e3

#-----------------------------------------------------------------------------
--#
# Run while loop until everyone in the simulation has died
ptime             <- system.time({
  while(
    # stopping criteria: each row in dt_next_event represents
    # the number of people still alive in the simulation
    nrow(l_params$dt_next_event) >0
  ){

    # Function updates next event for everyone who is still alive
    l_params$dt_next_event <- sim_next_event(l_params)

    # Warning:
    if(unique(length(l_params$sim_next_event$Event_num)) > max_num_events){
      warning(
        paste0("Within a simulation time-horizon of"
```

```
               ,l_params$time_horizon[2]-l_params$time_horizon[1], "years,
                the max number of events (i.e., transitions between health
                states) for at least one person in the simulation has
                exceeded", max_num_events,".Reconsider if the simulation
                should continue since these dynamics might not reflect
                what you intended."
        ))
    }
  }
})[3]
ptime/60
ptime

#View(dt_event_history)

# ------------------------------------------------------------------ #
# Trace Matrix ----
# ------------------------------------------------------------------ #

## A. Continuous time trace matrix ----
#* We want to recover the average state occupancy of the simulated cohort fro
m the individual
#* state transition trajectories. This mapping is encoded in the function `co
nt_time_trace_sick_sicker`
#* that returns a trace matrix. Note that in contrast to a discrete time trac
e matrix, where rows
#* index cycles, in the continuous time trace matrix, rows index events.

dt_CT_trace <- cont_time_trace_sick_sicker(dt_event_history = dt_event_histor
y,
                                           v_names_states   = l_params$v_stat
es,
                                           time_init        = l_params$time_h
orizon[1],
                                           proportions      = TRUE,
                                           rounding_factor  = 5,
                                           long            = F)

dt_CT_trace_long <- cont_time_trace_sick_sicker(dt_event_history = dt_event_h
istory,
                                                v_names_states   = l_params$v
_states,
                                                time_init        = l_params$t
ime_horizon[1],
                                                proportions      = TRUE,
```

```r
                                                rounding_factor  = 5,
                                                long             = T)
View(list(dt_CT_trace,dt_CT_trace_long))
### B.2 Discrete time approximation  ----
#* Many factors like sim sample size, large number of states, long time-horiz
ons,
#* can contribute to extremely large continuous time trace matrices,  `dt_CT_
trace`.
#* In those cases it might be more practical to use an approximation for task
s like plotting the trace.
#* The function `discr_time_trace_sick_sicker` provides such approximation by
finding the event times
#* closest to a grid of fixed times points supplied by the user. For example,
if we want to compare our DES implementation of the Sick Sicker model
#* with the discrete time, time dependent cSTM implementation we can supply t
he same time grid used for the cSTM and find
#* the closest evenT time to each time point on the grid.

# Discrete Time grid for a cSTM
cycle_length <- 1/12 # cycle length equal to one year (use 1/12 for monthly)
n_age_init   <- 25  # age at baseline
n_age_max    <- 100 # maximum age of follow up
n_cycles     <- (n_age_max - n_age_init)/cycle_length # time horizon, number
of cycles

v_DT_grid        <- seq(l_params$time_horizon[1],l_params$time_horizon[2], le
ngth.out = n_cycles)

dt_DT_trace      <- discr_time_trace_sick_sicker(dt_CT_trace     = dt_CT_trac
e ,
                                                 v_DT_grid   = v_DT_grid   ,
                                                 long        = F    )

dt_DT_trace_long <- discr_time_trace_sick_sicker(dt_CT_trace = dt_CT_trace ,
                                                 v_DT_grid   = v_DT_grid ,
                                                 long        = T     )
```

### 7. Auxiliary Module: Convergence Analysis

In this Module we illustrate how to assess the convergence of the state occupancy estimator as we increase the simulation sample size. State occupancy is recovered from the output of the DES by computing the proportion of simulated people in a given state at a given event time (Monte Carlo integration).

As we increase the simulation sample size, we expect the Monte Carlo standard error to shrink around the estimator of state occupancy. We simulate independent runs in batches of size `5e1` with varying simulation samples sizes, `1e2, 1e3, 1e4, 1e5`. We compute bootstrap confidence bounds (2.5 and 97.5 percentiles) around the median state occupancy over time.

```
#-----------------------------------------------------------------------------
--#
#* To program this tutorial we used:
#* R version 4.2.1 (2022-06-23)
#* Platform: aarch64-apple-darwin20 (64-bit)
#* Running under: Mac OS 12.5
#* RStudio: Version 2022.07.1+554
#-----------------------------------------------------------------------------
--#
# Description ----

#* We illustrate how to assess the convergence of the state occupancy estimat
or
#* as we increase the simulation sample size. State occupancy is recovered fr
om
#* the output of the DES by computing the proportion of simulated people in a
given state
#* at a given event time (Monte Carlo integration). As we increase the simula
tion sample size,
#* the Monte Carlo standard error shrinks  around the estimator of
#* state occupancy.

#-----------------------------------------------------------------------------
--#
# Initial setup ----
rm(list = ls())    # remove any variables in R's memory
gc()               # clean working memory
#-----------------------------------------------------------------------------
--#
## Load packages ----
library("data.table"  )
library("dplyr"       )
library("parallel"    )
library("foreach"     )
library("doParallel"  )
library("abind")
```

```r
## Load supplementary functions ----
source("R/DES_functions.R") # DES specific
source("R/Functions.R")     # General utility functions

#------------------------------------------------------------------------------
--#
# Initialize Parameters ----
#* We want to pass parameters seamlessly through our function sim_next_event(
),
#* for this we can declare initial parameters in a list. Here we pass them th
rough
#* a function `init_param` that will return such list `l_params` and also inc
lude the baseline data table
#* `dt_baseline` as an object of that list, now named `dt_next_event`.

l_params <- init_params(
  # Sim. params
  # Time horizon
  time_init         = 25    ,
  time_end          = 100   ,
  # Sim. sample size
  n_sim             = 1e2   ,

  # dt_baseline information
  ## initial age
  age_init          = 25    ,
  ## sex group
  Sex_grp           = "Total",
  ## initial state
  state_init        = "H"   ,

  # State Space
  v_states          = c("H",                                            # He
althy (H)
                        "S1",                                           # Si
ck (S1)
                        "S2",                                           # Si
cker (S2)
                        "D"),                                           # De
ad (D)

  # Indexed Adjacency Matrix
  #* Zero entries indicate no transition between pairs of states,
  #* Nonzero positive entries index possible transitions in arbitrary order
  m_Tr_r            = matrix(c(0,1,0,2,
                               3,0,4,5,
                               0,0,0,6,
                               0,0,0,0),
                             nrow = 4, ncol = 4, byrow = TRUE,
```

```
                            dimnames = list(c("H", "S1","S2","D"),
                                            c("H", "S1","S2","D"))),
  # Transition Rates
  ## Mortality
  #* we specify subfolder in project to load Life Tables
  lifetable        = "Inputs/all_cause_mortality.Rda",
  ## Hazard Ratio while being S1
  hr_S1            =    3,
  ## Hazard Ratio while being S2
  hr_S2            =    10,

  ## Other transitions
  ## H  -> S1, constant rate ~Exp(r_HS1)
  r_HS1            =    0.15,
  ## S1 -> H,  constant rate ~Exp(r_S1H)
  r_S1H            =    0.5,
  ## S1 -> S2, state-residency time dependent ~ Weibull(r_S1S2_scale_ph, r_S1
S2_shape)
  ##* PH parameterization
  r_S1S2_scale_ph  =    0.08,
  r_S1S2_shape     =    1.1,

  # Strategy
  #* Can be any of "SoC", "A", "B", "AB".
  #* Only B and AB, affect transitions rates,
  strategy         =    "SoC",

  # Tx Effects that affect state-transitions
  #* Effectiveness of strategy B hazard ratio
  #* of becoming Sicker when Sick under treatment B
  hr_S1S2_trtB = 0.6
)
#-----------------------------------------------------------------------------
--#
# -------------------------------- #
# Distributed Implementation ------
#  ------------------------------- #

## Cluster Setup ----

# Num. of Cores
no_cores_all  <- parallel::detectCores()-1      # recommended when not sharin
g computing resources
# cores_90pct   <- as.integer(.90 * no_cores_all) # aggressive  when shared r
esources
cores_85pct   <- as.integer(.85 * no_cores_all) # aggressive  when shared res
ources
# cores_75pct   <- as.integer(.75 * no_cores_all) # aggressive  when shared r
esources
```

```
# cores_50pct   <- as.integer(.5  * no_cores_all) # recommended when shared r
esources
# cores_25pct   <- as.integer(.25 * no_cores_all) # recommended when shared r
esources

os_info <- Sys.info()
os <- os_info[1]
no_cores <- cores_85pct
if (os == "Darwin") { # Apple Kernel
  # Initialize cluster object
  cl <- parallel::makeForkCluster(no_cores)
  # Register clusters
  doParallel::registerDoParallel(cl)
}
if (os == "Linux") {
  # Register clusters
  doMC::registerDoMC(cores = no_cores)
}
if (os == "Windows") {
  # Initialize cluster object
  cl <- parallel::makeCluster(no_cores)
  # Register clusters
  doParallel::registerDoParallel(cl)
}

## Run distributed implementation ----

# Set seed for reproducibility
n_seed            <- 2
set.seed(n_seed)

# batch and sim sample size
v_n_sim     <- c(1e2,1e3,1e4,1e5)   # iterate over
v_n_batch   <- c(5e1,1e2,1e3)       # fix one
k <- 5#v_n_batch[1]

#----------------------------------------------------------------------------
--#
#----------------------------------------------------------------------------
--#

# WARNING: DO NOT RUN THE NEXT CHUNK UNLESS YOU HAVE ENOUGH COMPUTING RESOURC
ES
# Should be fine with M1 Apple or similar
#----------------------------------------------------------------------------
--#
#----------------------------------------------------------------------------
--#
```

```
#* Function  `run_parallel_model` implements a distributed version of the DES
#* k times for a given sim sample size,  calls function `obtain_model_grid_tr
ace` to
#* compute the trace matrix approximation in discrete time k times.
#* Trace Matrices  are stored in an array `a_grid_trace_byState`.

#* Function `batch_summary` summarizes the k trace matrices and returns diffe
rent quantiles, mean and SE of
#* state occupancy for a give simulation sample size.

# dir.create("analysis", showWarnings = FALSE) # Create directory if it doesn
't exist
# # ------------------ #
# ## run for n_sim 1e2
l_inputs                     <- init_params(n_sim = v_n_sim[1])
a_grid_k_5e1_b_size_1e2      <- run_parallel_model(model_fn = obtain_model_gr
id_trace, l_inputs = l_inputs, k=k)
#Summary
batch_summary_k_5e1_b_1e2    <- batch_summary(array_grid = a_grid_k_5e1_b_siz
e_1e2, k = k, n_sim = v_n_sim[1])

rm(a_grid_k_5e1_b_size_1e2)
# # ------------------ #
# ## run for  n_sim 1e3
l_inputs                     <- init_params(n_sim = v_n_sim[2])
a_grid_k_5e1_b_size_1e3      <- run_parallel_model(model_fn = obtain_model_gr
id_trace, l_inputs = l_inputs, k=k)
# Summary
batch_summary_k_5e1_b_1e3    <- batch_summary(array_grid = a_grid_k_5e1_b_siz
e_1e3, k = k, n_sim = v_n_sim[2])
rm(a_grid_k_5e1_b_size_1e3)
# # ------------------ #
# ## run for n_sim 1e5
l_inputs <- init_params(n_sim = v_n_sim[4])
a_grid_k_5e1_b_size_1e5      <- run_parallel_model(model_fn = obtain_model_gr
id_trace, l_inputs = l_inputs, k=k)
# Summary
batch_summary_k_5e1_b_1e5    <- batch_summary(array_grid = a_grid_k_5e1_b_siz
e_1e5, k = k, n_sim = v_n_sim[4])
rm(a_grid_k_5e1_b_size_1e5)

# STOP CLUSTER
stopCluster(cl)
#----------------------------------------------------------------------------
--#
#----------------------------------------------------------------------------
--#
```

```
# Summaries ----

batch_summary_all <- as.data.frame(rbind(batch_summary_k_5e1_b_1e2,
                                         batch_summary_k_5e1_b_1e3,
                                         batch_summary_k_5e1_b_1e5)) %>%
  mutate(
    Sim_n = ifelse(n_sim == 100, "Sim. sample size = 1e2",
                   ifelse(n_sim == 1000, "Sim. sample size = 1e3",
                          "Sim. sample size = 1e5"))

  ) %>% mutate( Sim_n = factor(Sim_n, levels = c("Sim. sample size = 1e2","Si
m. sample size = 1e3","Sim. sample size = 1e5") ),
                State = factor(State, levels = c("Healthy", "Sick","Sicker",
"Dead")))

table(batch_summary_all$n_sim)
```

#### 8. Reproducing Plots in the Manuscript

To reproduce the plots shown in the manuscript please refer to the GitHub repository. The folder titled 'Plotting_Modules' contains R scripts to reproduce the plots for all analysis shown in the manuscript. Note that specialized plotting dependencies are required. To guarantee an exact replication of the results shown in the manuscript, follow the instructions in the GitHub repository to install the version of dependencies that the developer used.
